# Mapping the plasma proteomic architecture of systemic lupus erythematosus

**DOI:** 10.1101/2025.11.14.25340226

**Authors:** Geoffrey H. D. Leung, Charlotte Bottomley, Norzawani Buang, Robert T. Maughan, Benjamin J. Whittle, Boroumand Zeidaabadi, Yun-Ju Huang, Tabitha Turner-Stokes, Marie Condon, Liz Lightstone, Tom Cairns, Marina Botto, Matthew C. Pickering, James E. Peters

**Affiliations:** Department of Immunology and Inflammation, Imperial College London, London, UK; School of Medicine, Chang Gung University and Division of Rheumatology, Allergy and Immunology, Chang Gung Memorial Hospital, Taoyuan, Taiwan; Imperial Lupus Centre, Imperial College Healthcare NHS Trust, London, UK

## Abstract

Systemic lupus erythematosus (SLE) is a systemic autoimmune disease characterised by autoantibodies to nuclear antigens. SLE is highly heterogeneous, both clinically and immunologically, yet the molecular basis underlying this remains incompletely understood. To address this, we profiled the plasma proteome in 260 SLE patients and 86 healthy volunteers (HVs) using the SomaScan v4.1 platform, quantifying 7,288 analytes corresponding to 6,595 unique proteins. We identified 215 proteins that were robustly differentially abundant between SLE patients and HVs in both discovery (n=207 SLE, n=45 HVs) and validation sets (n=53 SLE, n=41 HVs). Within-cases analyses identified 421 proteins associated with disease activity. Network-based clustering delineated correlated protein modules, including an interferon-associated module and a renal-associated module, each linked to distinct clinical features. Autoantibody-stratified analyses further uncovered distinct proteomic endotypes: anti-Sm positivity was associated with increased interferon-stimulated protein levels (e.g., MX1, ISG15, CXCL10) and reduced circulating small ribonuclear proteins, independent of disease activity. Anti-dsDNA antibodies were associated with elevated levels of CD40 ligand (CD40LG) and the neutrophil protease proteinase-3. Moreover, we identified an association between CD40LG and disease activity specific to the anti-dsDNA positive subgroup. Together, these data define plasma protein signatures of SLE and disease activity, highlight autoantibody-specific molecular phenotypes, and provide a basis for precision medicine.

**Graphical abstract:** 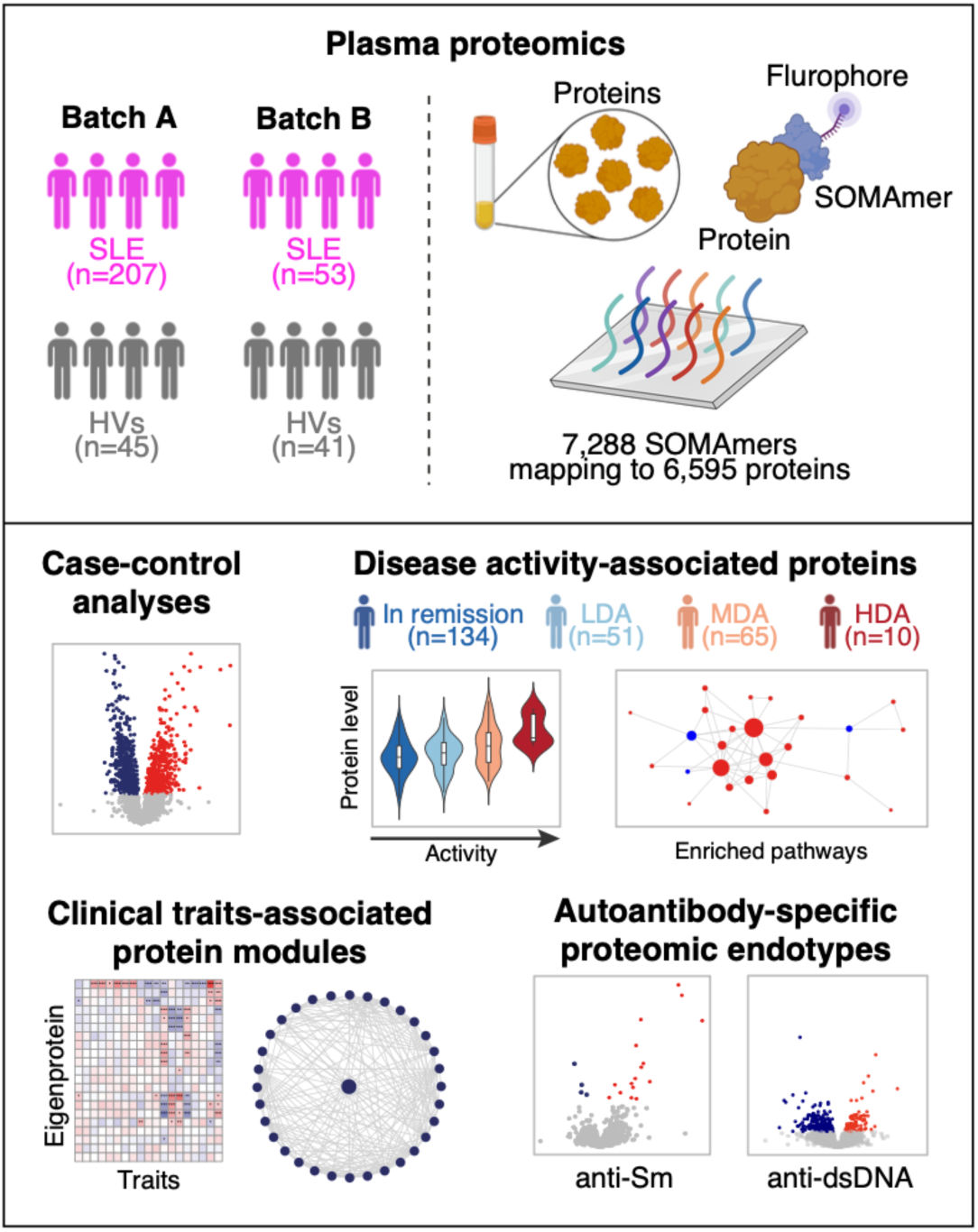

## Introduction

Systemic lupus erythematosus (SLE) is an autoimmune disease with complex pathogenesis involving dysregulation of both adaptive and innate immunity. A hallmark of SLE is the production of autoantibodies directed against nuclear antigens (anti-nuclear antibodies; ANA). Established pathogenic pathways include autoreactive B cells, the complement system and type 1 interferon signalling which lead to tissue inflammation and injury. Clinically SLE is highly heterogeneous, ranging from mild to organ- or life-threatening disease, and with variable patterns of organ involvement. There is also inter-patient variability in the targets of ANA. This heterogeneity presents major challenges for patient management, drug development, and trial design. There is a pressing need for a precision medicine approach, matching the right therapy to the right patient. Our ability to deliver such a tailored therapeutic strategy is hampered by insufficient understanding of the molecular basis of heterogeneity and we lack biomarkers that can identify patient subgroups who would benefit from pharmacological targeting of specific pathways.

The advent of genomic technologies has enabled comprehensive assessment of various molecular domains in SLE such as the genome or transcriptome. Genome-wide association studies (GWAS) have elucidated genetic risk factors for SLE [1–3], while transcriptomic studies have revealed the dysregulated gene expression programmes that occur once disease is established [4–6]. A key finding from the latter was the identification of the interferon-stimulated gene (ISG) signature, a gene expression signature indicating stimulation by interferons, particularly type 1 interferons [4, 7–9]. By contrast, few studies have investigated the proteome in SLE [10–12]. Proteins are the effector molecules of biology and the targets of most drugs. In addition, circulating proteins (measurable in plasma or serum) can provide useful and tractable biomarkers suitable for measurement in clinical practice. Thus, evaluation of proteomic changes in SLE could be valuable both for understanding pathogenesis and for translational research [13]. Advances in proteomic technologies now allow for highly multiplexed protein measurements. Here we performed plasma proteomic profiling using the SomaScan platform, measuring 7,288 protein analytes. Our data identify differentially abundant proteins in SLE versus HVs, protein signatures of active disease, and modules of correlated proteins associated with clinical traits. Moreover, we identify auto-antibody specific proteomic endotypes, with distinct proteomic patterns correlated with Smith and dsDNA auto-antibody positivity. These findings provide a basis for precision medicine approaches in SLE.

## Results

### Plasma proteomic profiles of SLE

We recruited 260 patients with SLE and 86 healthy volunteers (HVs). The cohort encompassed a wide range of disease duration and disease activity (**Table 1**). Seventy patients (28%) had anti-double-stranded DNA (dsDNA) autoantibodies, 193 (75%) anti-Smith (Sm) autoantibodies, and 186 (72%) had anti-ribonucleoprotein (RNP) autoantibodies (**Table 1**). Reduced complement C3 (<0.70 g/L) was observed in 14% of samples and reduced complement C4 (<0.16 g/L) in 26%. One hundred and fifty-eight patients (61%) had a history of previous lupus nephritis (LN) **(Supplementary Data 1)**, but only 10 patients had recent LN (defined as blood sampling within 3 months of a renal biopsy showing LN). Proteomic measurements were performed in two batches. After quality control, 260 SLE and 86 HV samples were available for analysis. Batch A (“discovery set”) consisted of 207 SLE samples and 45 HV samples and batch B (“validation set”) consisted of 53 SLE samples and 41 HV samples.

**Table 1.** Characteristics of study participants. P-value for differences in sex distribution was calculated using chi-squared test; P-values for differences in age were calculated using Student’s t-test. *Recent nephritis defined as blood sample taken within 3 months of a renal biopsy showing lupus nephritis. †Anti-dsDNA antibodies and complement levels measured at the time of research sample. ‡Differences in N reflect individuals with missing data for autoantibody status.

|  |  | **SLE** | **Healthy** | **P value** |
| --- | --- | --- | --- | --- |
| **N, Samples [%]** |  | 260 | 86 |  |
|  | Female | 229 [88.1%] | 58 [67.4%] | <0.0001 |
|  | Male | 31 [11.9%] | 28 [32.6%] |  |
| **Median age, Years [IQR]** |  | 43.9 [34.8, 55.4] | 39.5 [30.0, 49.1] | 0.001 |
|  | Female | 44.0 [34.6, 55.5] | 39.6 [30.3, 48.1] | 0.002 |
|  | Male | 39.9 [35.3, 51.9] | 39.4 [30.6, 50.4] | 0.271 |
| **N, Disease Activity [%]** |  | 260 | - |  |
|  | In remission | 134 [51.5%] | - |  |
|  | Low | 51 [19.6%] | - |  |
|  | Moderate | 65 [25%] | - |  |
|  | High | 10 [3.8%] | - |  |
| **Median disease duration, Years [IQR]** |  | 12.0 [5.5, 19.6] | - |  |
| **N, Lupus nephritis [%]** |  |  |  |  |
|  | Never | 92 [35.4%] |  |  |
|  | Previous | 158 [60.8%] | - |  |
|  | Recent* | 10 [3.8%] | - |  |
| **Laboratory parameters** |  |  |  |  |
|  | **C3 level (g/L, Median [IQR])†** | 1.03 [0.85, 1.20] | - |  |
|  | N, Low C3 (<0.7g/L) [%] | 36 [13.8%] | - |  |
|  | **C4 level (g/L, Median [IQR])†** | 0.21 [0.15, 0.28] | - |  |
|  | N, Low C4 (<0.16g/L) [%] | 68 [26.2%] | - |  |
|  | **N, Anti-Sm status‡ [%]** | 256 | - |  |
|  | Negative | 193 [75.4%] | - |  |
|  | Positive | 63 [24.6%] | - |  |
|  | **N, Anti-RNP status [%]** | 260 | - |  |
|  | Negative | 186 [71.5%] | - |  |
|  | Positive | 74 [28.5%] | - |  |
|  | **N, Anti-dsDNA status†‡ [%]** | 250 | - |  |
|  | Negative (0-9 IUperML) | 140 [56%] | - |  |
|  | Borderline (10-20 IUperML) | 40 [16%] | - |  |
|  | Positive (>20 IUperML) | 70 [28%] | - |  |

To identify proteomic signatures associated with SLE, we performed differential protein abundance testing of SLE versus HVs. For readability, where reporting the number of associations we use the term ‘protein’ to refer to the protein target of each SOMAmer, but provide further detail of SOMAmer to protein mapping in the Supplementary Data. In Batch A (n=207 SLE, n=45 HVs), we identified 1,821 proteins associated with SLE (1,065 upregulated, 756 downregulated; FDR <0.05) (**Figure 1A**, **Supplementary Figure 1A, Supplementary Data 2**). In batch B (53 SLE, 41 HVs), we identified 428 differentially abundant proteins (268 upregulated, 160 downregulated; FDR <0.05) **(Supplementary Figure 1B, Supplementary Data 3)**. 215 of the 1,821 significant associations from batch A replicated in batch B at FDR <0.05 **(Figure 1A, Supplementary Data 4)**. The estimated effect sizes for these replicated proteins were strongly correlated between the two batches (Pearson’s r 0.94) (**Figure 1B**). We then performed a meta-analysis of the two batches to obtain an overall estimate of the effect sizes for the replicated proteins (**Figure 1C. Supplementary Data 4**). Notably, there was marked upregulation of interferon-stimulated proteins (ISPs) in SLE, including ISG15, MX1, STAT1, B2M, DDX58, IFIT3, CXCL11 **(Figure 1D)**. Other strongly upregulated proteins included TNFRSF1B (TNF-R2), VSIG4 (the receptor for complement C3b and iC3b) and the complement fragment C3d. The most downregulated protein was CRISPLD2, a secreted LPS-binding glycoprotein that dampens TLR4-mediated cytokine responses [14]. Gene Set Enrichment Analysis (GSEA) identified 8 pathways that were significantly enriched in both batches, including multiple terms reflecting interferon signalling, as well as SRP-mediated cotranslational protein targeting, and downregulated PI3K/AKT signalling **(Figure 1E, Supplementary Data 5).**

**Figure 1.**
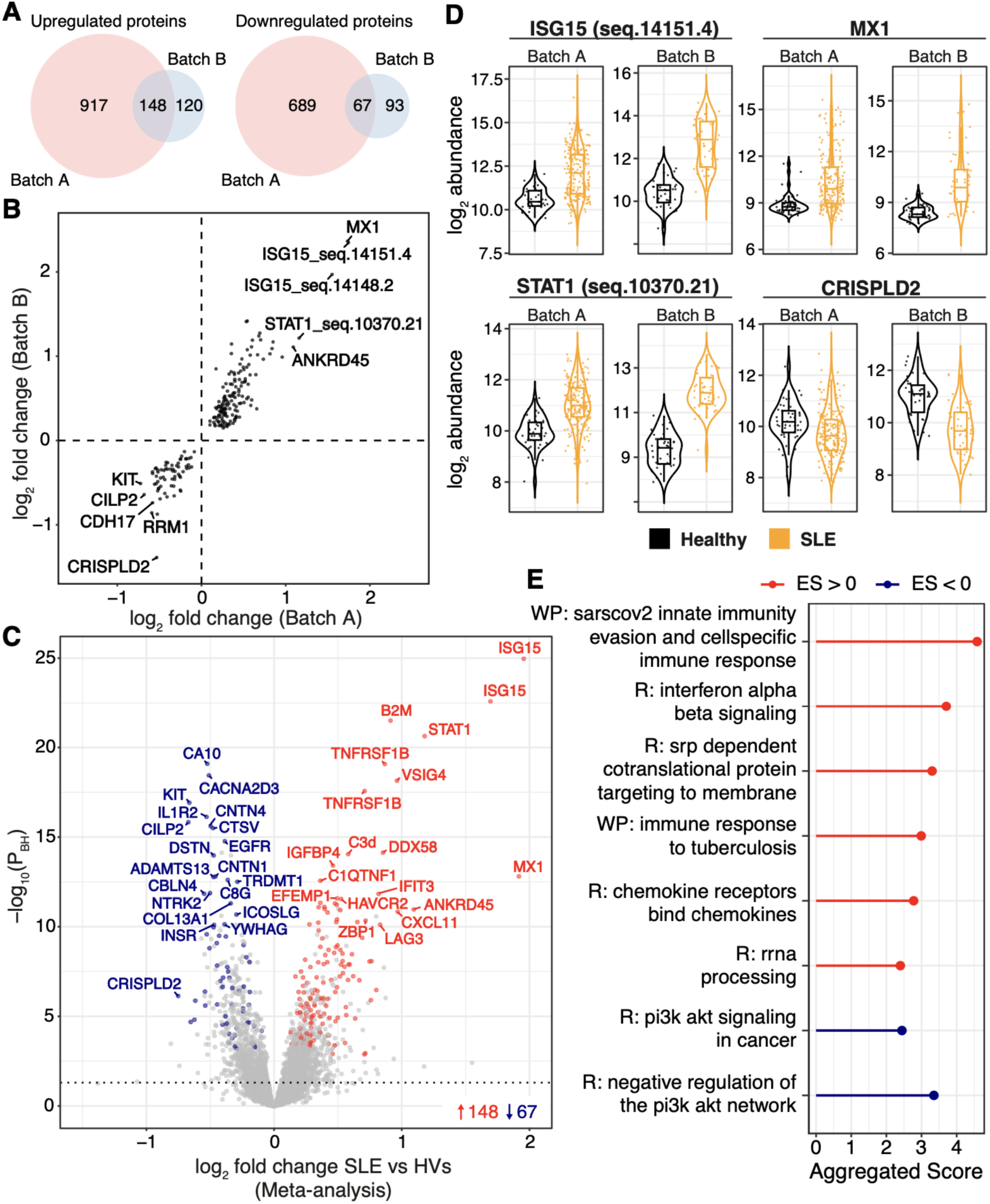
Differentially abundant proteins and enriched pathways in SLE versus healthy volunteers. **A)** Venn diagrams of the number of significantly (FDR <0.05) upregulated (left) and downregulated (right) proteins in SLE versus HVs in batch A (n=207 SLE, n=45 HVs) and B (n=53 SLE, n=41 HVs) data. **B)** Comparison of effect size estimates (covariate-adjusted log_2_ fold change) for validated proteins between Batch A and Batch B. Each point represents the protein target of a SOMAmer. 215 protein targets which were significantly differentially abundant (FDR <0.05) in both batch A and batch B are shown. The top 5 upregulated and downregulated protein targets (ranked by meta-analysed effect sizes) are annotated. **C)** Volcano plot showing the results from meta-analysis of Batches A and B. Coloured points represent protein targets replicated in both batches (FDR <0.05). Red: upregulated, blue: downregulated, grey: not replicated or non-significant. **D)** Examples of most significant differentially abundant proteins. Each point represents a sample. Orange: SLE, Black: HVs. **E)** Pathways enriched (FDR <0.05) in both batches. Aggregated score was calculated using robust rank aggregation. Red: upregulated pathways. Blue: downregulated pathways. R, Reactome; WP, WikiPathways; ES, Effect size.

### Plasma proteomic signatures of lupus disease activity

Samples were obtained from patients with a range of disease activities; 134 patients were in clinical remission, 51 had low disease activity (LDA), 65 had moderate disease activity (MDA), and 10 had high disease activity (HDA) (definitions in **Methods**). We tested for proteomic associations with SLE disease activity in SLE coded as ordinal variable. To increase power, we analysed both Batch A and B together, adjusting for batch. 421 proteins were significantly associated (FDR <0.05) with disease activity (**Figure 2A, Supplementary Data 6**). As a sensitivity analysis, we performed Spearman’s correlation analysis between SLEDAI and each protein, which showed similar results (**Supplementary Figure 2**).

**Figure 2.**
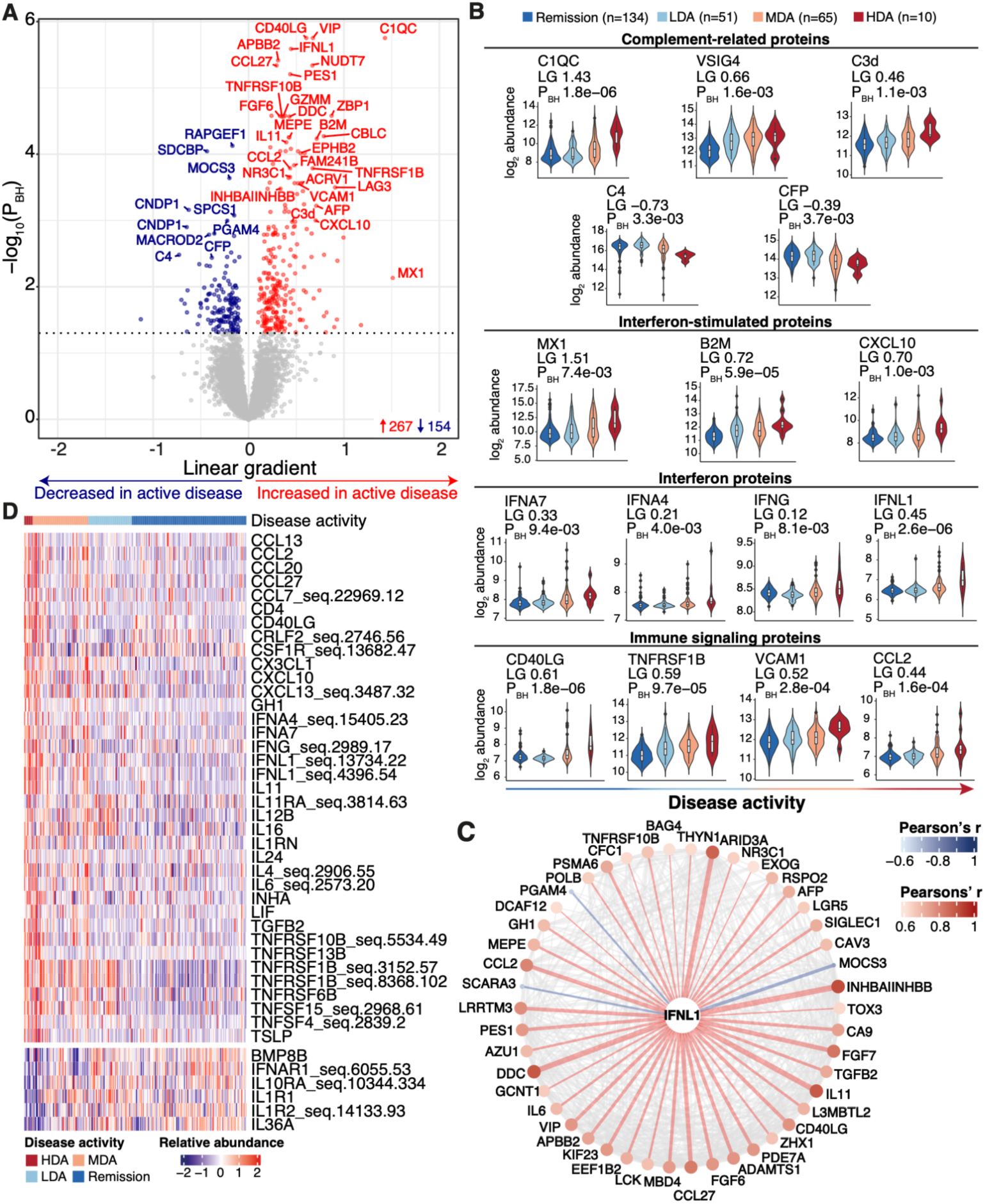
Proteins associated with SLE disease activity. n=260 SLE samples. **A)** Volcano plot. P_BH_ Benjamini-Hochberg-adjusted P-value (linear regression). Linear gradient shows the magnitude and direction of change in the protein level with increasing disease activity. Dotted line = 5% FDR. Each point represents a protein. Red: significant positive association with active disease, blue: negative association, grey: non-significant. **B)** Examples of disease-associated complement-related, interferon-pathway and immune signalling proteins. LG = Linear gradient. Unique SOMAmer identifiers for displayed proteins where the SomaScan has >1 SOMAmer are: B2M (seq.3485.28), IFNA4 (seq.15405.23), IFNG (seq.2989.17), TNFRSF1B (seq.3152.57), and VCAM1 (seq.2967.8). **C)** Disease activity-associated proteins correlated with IFNL1. For clarity of visualisation, only the proteins associated with disease activity at 1% FDR are displayed. A detailed list of proteins correlated with IFNL1 is provided in Supplementary Data 8. Edges represent pairwise protein correlations with |Pearson’s r|>0.6. Node size, colour, and edge thickness indicate the strength of correlation (Pearson’s r) with IFNL1. Edge colour denotes correlation direction: blue for negative and red for positive. **D)** Heatmap of significant (FDR <0.05) disease activity-associated proteins in the KEGG Cytokine-Cytokine Receptor Interaction pathway. Protein levels were adjusted for sex and batch.

GSEA revealed significant enrichment of 90 pathways (**Supplementary Data 7**), the most significant of which were “immunoregulatory interactions between a lymphoid and a non-lymphoid cell” and “cytokine-cytokine receptor interaction”. The proteins most strongly associated with disease activity by p-value were CD40LG, VIP (vasoactive intestinal peptide) and C1QC (all P_BH_ 1.77×10^-6^). Active disease was also associated with other complement-related proteins, including increased C3d and VSIG4 and decreased C4 and CFP (properdin) (**Figure 2B**). ISPs (MX1, B2M, CXCL10) were positively associated with disease activity, with MX1 showing the greatest effect size. Levels of type I, II and III interferons were also associated with active disease, with IFNL1 (a type III interferon) showing the strongest effect. Evaluation of connections between IFNL1 and other disease activity-associated proteins revealed IFNL1 was co-correlated (|Pearson’s r| >0.6) with 75 other proteins (**Figure 2C, Supplementary Data 8**).

The SomaScan includes many proteins that are primarily intracellular. Their detection in plasma may reflect cell death and turnover. While such proteins may be useful biomarkers of disease activity, some are likely to be downstream read-outs of tissue inflammation and injury rather than upstream pathogenic drivers. In contrast, plasma proteins associated with disease activity that have a biological role in blood may be more promising therapeutic candidates. For example, cytokines and their receptors have proven to be important therapeutic targets in inflammatory diseases. We therefore extracted a list of proteins annotated in the KEGG ‘Cytokine–cytokine receptor interaction’ pathway and intersected this with the list of SLE activity-associated proteins (**Figure 2D**). In active SLE, there was upregulation of TNF superfamily and TNF receptor superfamily members (e.g., TNFSF4 (OX40L), TNFSF15 (TL1A), TNFRSF1B (TNFR2), TNFRSF10B (DR5)), chemokines (e.g., CCL2, CCL27, CXCL10, CXCL14), interleukins (e.g., IL6, IL11, IL24), and growth factors. In contrast, there was downregulation of anti-inflammatory proteins including IL10RA and the decoy receptor IL1R2.

### Associations with complement proteins

Given the strong links between complement proteins and disease activity in the literature and in our data, we further analysed proteomic associations with serum complement C3 and C4 measured in the clinical laboratory (n=260 SLE patients). Both C3 and C4 were also included on the SomaScan assay. There was very strong concordance of protein measurements between the SomaScan and the clinical laboratory for C4 (ρ 0.95 in Batch A, 0.91 in Batch B), and moderately strong correlation for C3 (ρ 0.61 in Batch A, 0.45 in Batch B) (**Supplementary Figures 3A-B**), providing validation of SomaScan measurements. In addition, various complement cleavage fragments were measured; correlations with intact C3 and C4 are presented in **Supplementary Figures 4-5**. Sixty protein targets were correlated with clinical laboratory measured C3 (|ρ| > 0.3, FDR < 0.05), **Supplementary Data 9**) including 21 associated with disease activity (**Supplementary Figures 6A-B**). Sixty-two proteins were significantly correlated with C4 (**Supplementary Data 10**), 13 of which were disease activity associated (**Supplementary Figure 6C**). In total, 24 protein targets were associated with disease activity and levels of C3 and/or C4 (**Supplementary Figure 6D**). Proteins associated with higher disease activity and lower C3/C4 included CD40LG, interferon-stimulated proteins (e.g. ISG15, IFIT3, LAG3), and C1QC, while IL36A, complement-related proteins (CFP (properdin), FCN1 (ficolin) and C2) and IFNAR1 showed the opposite relationships. The latter may reflect compensatory downregulation in the context of chronic interferon stimulation.

### Exploratory Longitudinal Analysis

For 3 patients, longitudinal samples taken at 4 or more timepoints were available. The limited sample size precluded formal biomarker analysis, but we used these data to further explore 7 selected proteins (CXCL10, CCL2, CD40LG, HAVCR2, C3d, MX1, B2M) that we had identified as disease activity-associated in the cross-sectional analysis (see **Methods)**. The levels of these proteins generally mirrored disease activity over time, showing positive longitudinal correlations with SLEDAI-2K and anti-dsDNA, and negative correlations with C3/C4 (**Supplementary Figures 7A-B**). CD40LG strongly correlated with anti-dsDNA titres, in keeping with its role in B cell activation (**Supplementary Figure 7C**). Longitudinal changes in C3d were inversely correlated with clinically measured C3 (**Supplementary Figure 7D**), in keeping with the known biology of the complement pathway activation whereby cleavage of C3 reduces the amount of intact C3 and increases C3d. Several proteins returned to normal levels (defined based on HV samples) as disease activity declined, though the levels of the ISPs CXCL10, MX1 and B2M failed to fully normalise, suggesting persistent interferon pathway activation (**Supplementary Figure 7A)**.

### Network analysis reveals protein modules associated with clinical phenotype

We used weighted gene co-expression network analysis (WGCNA) to construct a protein-protein correlation network, identifying 21 protein modules. We then tested for associations between each module and traits including SLEDAI-2K score, ISG score (measured by quantitative polymerase chain reaction, qPCR) and clinical laboratory parameters. The red module, consisting of 35 proteins, was significantly correlated with multiple clinical traits, with positive correlation with SLEDAI-2K score, and multiple autoantibodies, and was negatively correlated with neutrophil and lymphocyte count and levels of haemoglobin, albumin, and complement C3 and C4 (**Figures 3A-E, Supplementary Figures 8A-G**). Notably, this module showed strongest correlation with ISG score (ρ 0.75, P_BH_ 2.4×10^-29^; **Figure 3B**), and pathway analysis showed enrichment for interferon signalling pathways (**Figure 3F**). ISG15 (measured by SOMAmer seq.14148.2) was the hub protein in this IFN-associated module (**Figure 3G, Supplementary Data 11**). ISG15 is an ISP and the module contained other ISPs (including CXCL10, CXCL11, DDX58, MX1, STAT1) and also IFNL1. While red module membership was dominated by interferon-related proteins, the module also contained other proteins including the complement component C1QC, the membrane protein APOL2, and LAG3. These protein network correlations may suggest interplay between C1Q, interferon pathways, and immune signalling. Specifically, LAG3 is a negative regulator of T cell responses, thus suggesting that interferon stimulation (indicated by upregulation of ISPs) may correlate with T cell inhibition.

**Figure 3.**
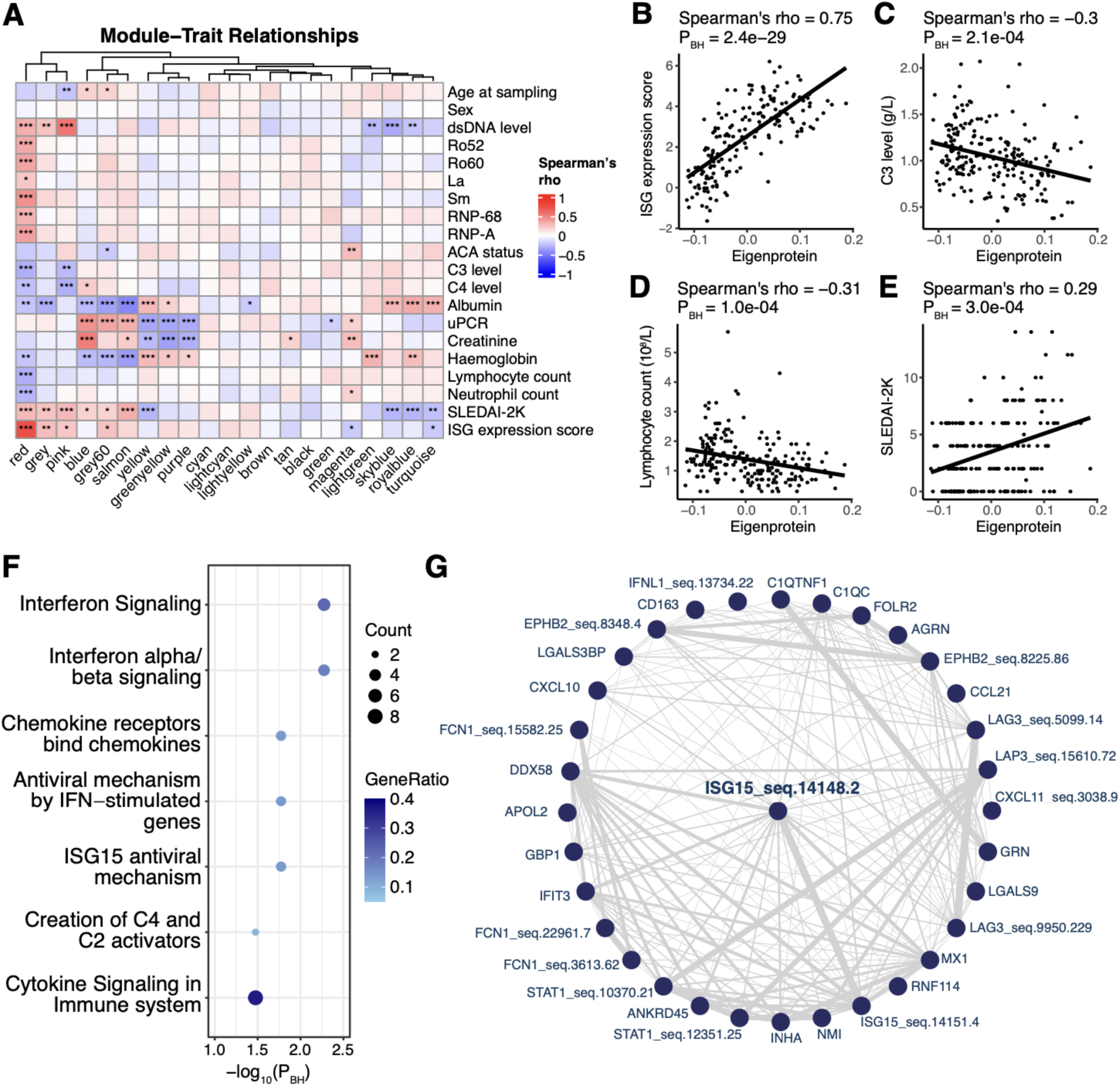
Network analysis identifies protein modules correlated with clinical traits. **A)** Network constructed using SLE samples (n=207). Correlation between each module and clinical traits. Asterixes represent BH-adjusted P values: * < 0.05, ** < 0.01, ***< 0.001. Colour gradient represents Spearman’s ρ. **B-E)** Relationships between the eigenprotein values of the red module and clinical traits: **(B)** ISG expression score, **(C)** C3 level, **(D)** lymphocyte count, **(E)** SLEDAI-2K. Each point represents a sample. Solid line indicates best fit from linear regression. **F)** Pathways enriched for proteins in the red module. **G)** Network of protein members of the red module. Edges represent the strength of similarity between each pair of proteins. ISG15 (measured by SOMAmer seq.14148.2) was the module hub protein.

The blue module, which had the second-highest number of clinical associations, was positively correlated with creatinine (ρ 0.52, P_BH_ 7.1×10^-14^) and proteinuria (ρ 0.45, P_BH_ 5.4×10^-10^) and negatively correlated with albumin **(Figures 3A, 4A-B)**, indicating a link to renal dysfunction. Consistent with this, the module contained Cystatin C (CST3, an accurate marker of glomerular filtration rate; **Figure 4C**), Osteopontin (SPP1, a biomarker of kidney injury), and Parathyroid hormone (PTH which is dysregulated in renal impairment); (**Figures 4C-D, Supplementary Data 12**). Other renal-related proteins included Ephrin and Ephrin receptors, which are associated with glomerular podocyte survival, angiogenic modelling and responses to renal injury [15, 16]. Comprising 611 proteins, this renal-associated module was enriched for extracellular matrix-related pathways, possibly reflecting renal scarring (**Figure 4E**). Moreover, the module eigenprotein was significantly higher in patients with a history of LN, particularly those with recent LN, compared with those who never had LN **(Figure 4F)**.

**Figure 4.**
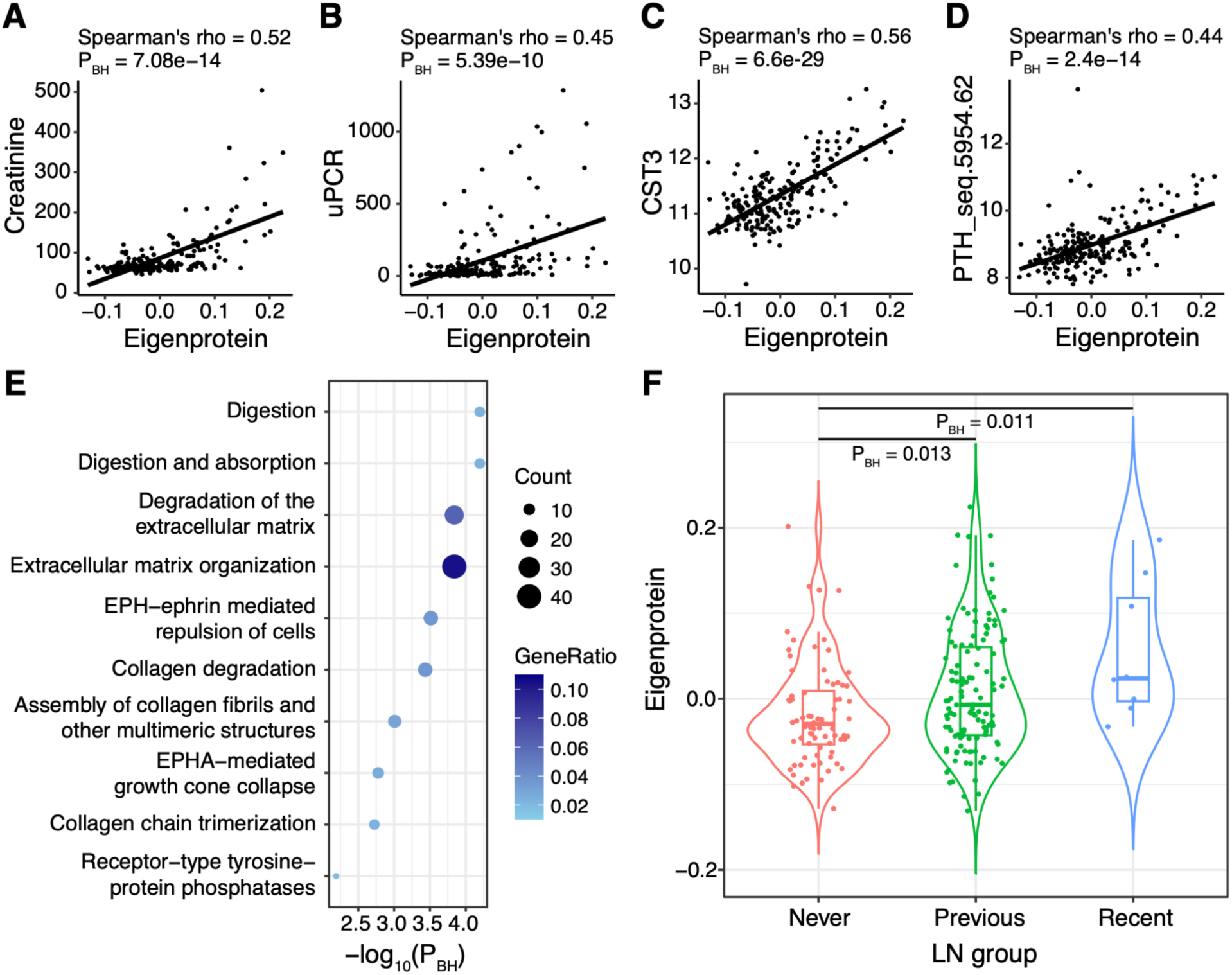
Renal-associated protein module. **A-D)** Relationships between the eigenprotein values of the blue module and renal markers: **(A)** Creatinine, **(B)** uPCR, **(C)** cystatin C (CST3), **(D)** parathyroid hormone (PTH). Each point represents an SLE sample. Solid line represents the line of best fit from linear regression. **E)** Pathways enriched for proteins involved in the blue module. **F)** Eigenprotein values of the blue module according to lupus nephritis (LN) status. n=207 SLE samples.

### Autoantibody-specific proteomic endotypes

Anti-Sm autoantibodies are highly specific for SLE and are associated with worse prognosis [17, 18]. To identify a proteomic signature associated with anti-Sm autoantibodies, we compared the abundance levels of each protein between anti-Sm(+) (n=63) and anti-Sm(-) (n=193) patients. Eighty-three proteins were differentially abundant, with 63 proteins upregulated and 20 downregulated in the anti-Sm(+) samples (**Figure 5A**). We considered the possibility that disease activity might be confounded with anti-Sm autoantibody positivity, since patients with Sm antibodies tend to have a worse prognosis and that some of the Sm-associated proteins were associated with disease activity in our previous analyses. We therefore performed multiple linear regression, controlling for disease activity. This revealed 54 proteins associated with anti-Sm antibodies independent of disease activity (43 proteins positively and 11 negatively correlated; **Figure 5B, Supplementary Data 13**). Visualisation of abundance of these proteins against anti-Sm status stratified by disease activity confirmed that the association with Sm antibodies was independent of disease activity (**Figure 5C, Supplementary Figure 9**). Pathway enrichment analysis revealed Sm-associated proteins were enriched for SARS-Cov-2-related, interferon signalling, snRNP assembly, and RNA processing pathway terms (**Figure 5D**). These pathway enrichment terms reflected increases in ISPs and reductions of small nuclear ribonucleoproteins (snRNPs) (**Figure 5E**). ISPs increased in Sm+ SLE included ISG15 (SOMAmer seq.14148.2), MX1, CXCL10, GBP1, DDX58, CXCL11, APOL2, STAT1, LGALS9, EPHB2, and LAG3. Since small nuclear ribonucleoproteins (snRNPs) are the targets of anti-Sm antibodies, it is possible that the reduction in circulating snRNPs reflects autoantibody-mediated depletion.

**Figure 5.**
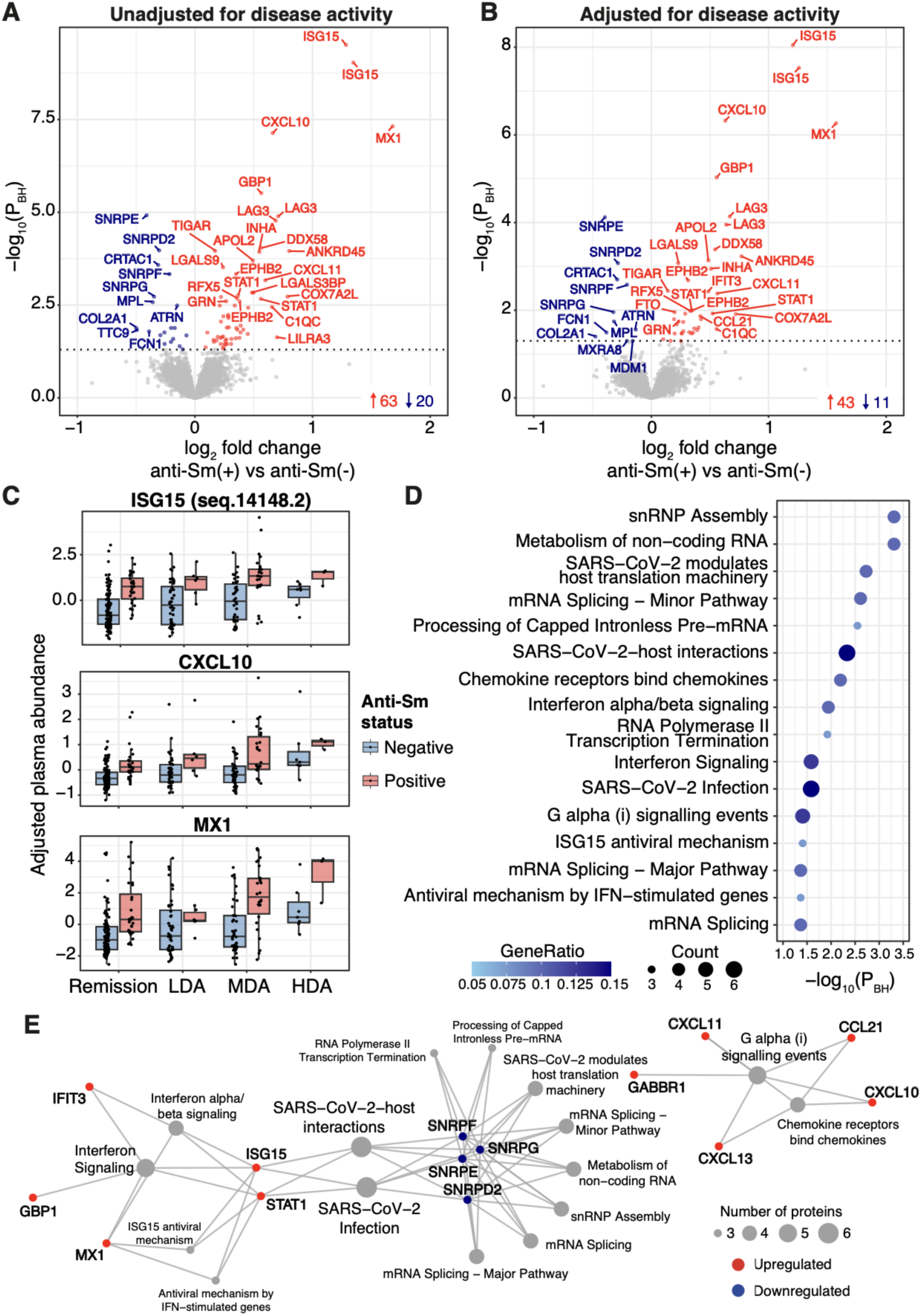
Proteins associated with anti-Sm autoantibodies. **A)** Proteins differentially abundant in anti-Sm positive (n=63) versus anti-Sm negative SLE (n=193), without adjustment for disease activity. Each point represents a protein. Red: significantly upregulated, blue: significantly downregulated, grey: non-significant. **B)** As for **A,** but after adjustment for disease activity. **C)** Interferon-stimulated proteins are associated with Sm antibodies independent of disease activity. Displayed protein levels have been adjusted for batch and sex. **D)** Significantly enriched pathways for proteins associated with anti-Sm antibodies after adjustment for disease activity. **E)** Connections between proteins associated with anti-Sm antibodies status independent of disease activity and the enriched pathways. Node size (for pathway terms) represents the number of proteins associated with the pathway. Red: upregulated proteins. Blue: downregulated proteins.

We then analysed the associations between proteins and anti-dsDNA antibodies. 874 proteins were associated with anti-dsDNA antibody positivity (**Figure 6A**). After adjusting for a modified disease activity score (which excluded dsDNA antibody status), 374 proteins remained associated with dsDNA antibody status (**Figures 6B-C, Supplementary Data 14**). The most upregulated proteins included CD40LG (CD40 ligand), a costimulatory molecule which activates B and T cells and promotes inflammatory responses [19], ARID3A (a DNA-binding protein), IL11, the neutrophil protease proteinase-3 (PRTN3), and RPS7. Proteins reduced in dsDNA antibody positive SLE included CFP, ITGAV|ITGB3 (the vitronectin receptor, an integrin heterodimer), L1CAM, BPIFB1, and C2. Unlike for Sm autoantibody status, the association of ISPs with anti-dsDNA antibody positivity became non-significant after adjustment for disease activity. As a sensitivity analysis, Spearman’s rank correlation was performed to test for correlations between the levels of anti-dsDNA autoantibodies (as a continuous trait) and each protein (**Supplementary Figure 10**). In this sensitivity analysis, CD40LG, PRTN3 and ARID3A were again among the strongest associations. Intriguingly, visualisation of CD40LG association with disease activity stratified by antibody status showed this relationship was restricted to the dsDNA antibody positive subgroup (**Figure 6D**; dsDNA+: P 4.2×10^-3^, beta 0.07; dsDNA-: P 0.52, neta −0.005). Formal interaction analysis confirmed this effect (P 6.7×10^-5^).

**Figure 6.**
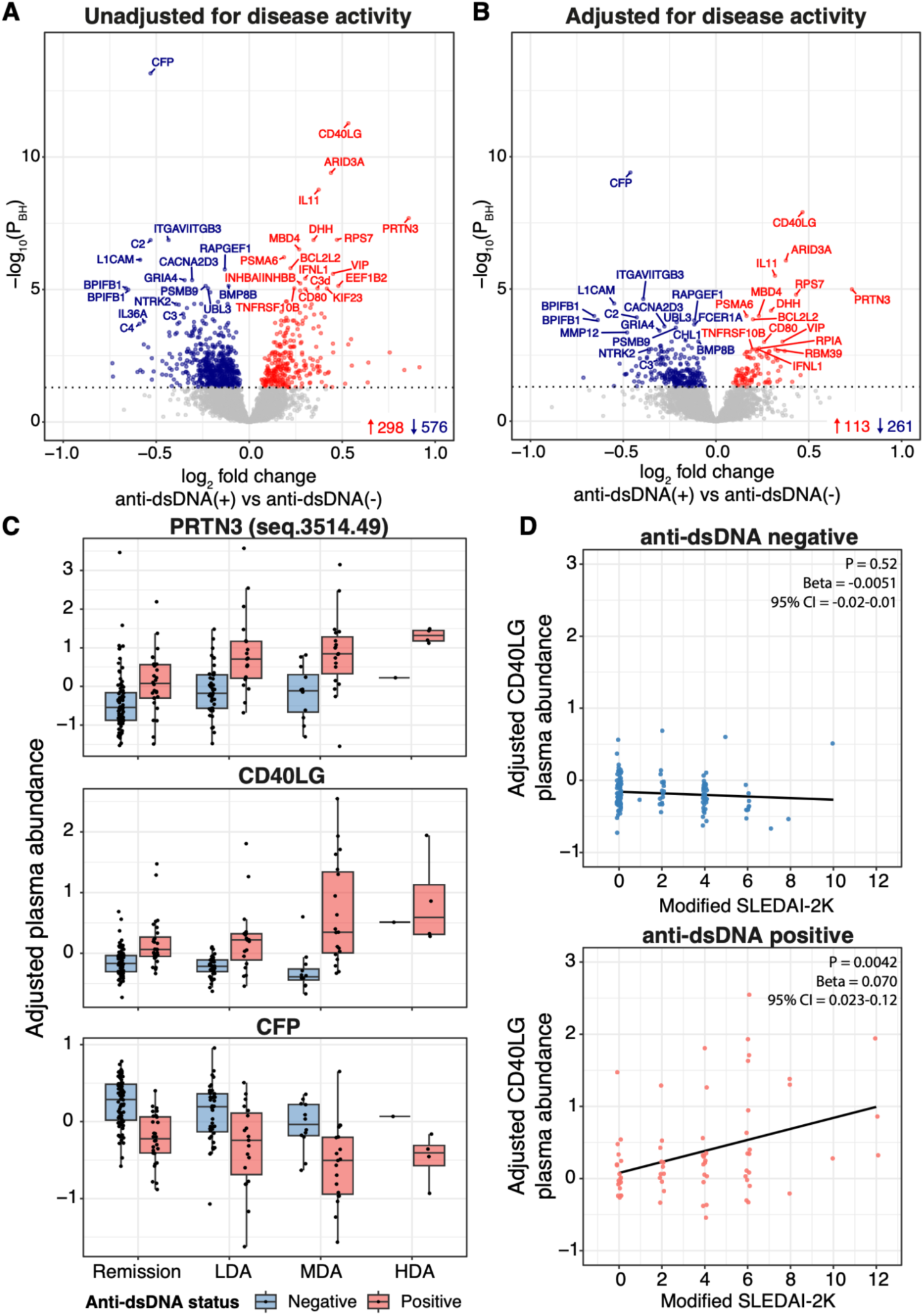
Proteins associated with anti-dsDNA autoantibodies. **A)** Proteins differentially abundant in anti-dsDNA positive (n=70) and anti-dsDNA negative (n=140) SLE samples, without adjustment for disease activity. Each point represents a protein. Red: significantly upregulated, blue: significantly downregulated, grey: non-significant. **B)** As for **A**, but after adjustment for disease activity. **C)** Examples of proteins associated with anti-dsDNA antibodies independent of disease activity. Protein levels in ant-dsDNA antibody positive and negative samples are shown stratified by disease activity. **D)** The association of plasma CD40LG with disease activity is restricted to anti-dsDNA positive individuals. SLEDAI-2K modified to exclude anti-dsDNA status from the score. Each point represents an SLE sample. Solid line represents the line of best fit from linear regression. Protein levels in **C** and **D** have been adjusted for batch and sex.

### Comparison with anifrolumab-induced proteomic changes in TULIP-1 trial

Given the prominence of interferon pathways in proteomic signatures in our data, we examined to what extent these signatures might be reversible with anti-type 1 interferon receptor therapy through comparison with an external dataset from the study of Baker et al which analysed 169 proteins in SLE pre- and post-anifrolumab in the TULIP-1 trial [20] using Olink and Simoa immunoassays. 158 of these proteins were measured in our SomaScan dataset (reflecting 228 SOMAmers since some proteins were measured by multiple SOMAmers). Across these 158 proteins, log_2_ fold changes from our SLE vs HV comparison were negatively correlated with anifrolumab-vs-placebo changes at week 52 in TULIP-1; (r −0.43; **Supplementary Figure 11A**). Similarly, protein associations with disease activity in our study were inversely correlated with changes following anifrolumab treatment (r −0.52; **Supplementary Figure 11B**). Of the proteins measured in both studies, 30 were associated with disease activity in our study, with 13 significantly modulated by anifrolumab treatment. These data underscore the centrality of the interferon response in plasma proteomic signatures of SLE. However, some disease activity-associated proteins were not modulated by anifrolumab, including CD40LG, indicating interferon-independent disease-activity related pathways (**Supplementary Figure 11C**).

## Discussion

We performed wide-angled proteomic profiling in lupus, measuring 7,288 analytes reflecting 6,595 unique proteins, representing the most comprehensive assessment of the plasma proteome to date. Proteins differentially abundant in SLE vs healthy volunteers and proteins associated with disease activity reflected known pathogenic pathways in SLE, including dysregulation of adaptive immunity, complement pathways, interferon and TLR signalling, validating the utility of plasma proteomics in uncovering underlying disease mechanisms.

Notably, there was strong upregulation of protein products of the ISG signature in SLE and this was more pronounced in active disease. The lupus ISG was first described in transcriptomic microarray studies of immune cells (whole blood or PBMCs) [4, 7, 21]. Here, we demonstrate that a similar ‘ISP’ signature is present at the protein level in the extracellular space in blood. Few broad-capture plasma proteomic studies have been conducted in SLE. One prior study used an earlier SomaScan version (1,129 proteins) identified 4 interferon-stimulated proteins (EPHB2, LAG3, CXCL13, and CXCL10) associated with SLE[10]. In our study, all of these proteins were significantly upregulated in SLE samples and with increasing disease activity, validating previous findings. A high ISG is associated with worse prognosis [9], but has not entered clinical practice. A plasma-based ISP rather than a gene expression assay might prove more tractable for clinical translation.

Among the most upregulated interferon-stimulated proteins in the present study were ISG15, MX1, STAT1, CXCL11, B2M and LAG3. LAG3 is an inhibitory receptor expressed on activated T cells. Its expression is increased in the context of chronic antigen exposure (e.g. cancer and chronic viral infection) and leads to T cell exhaustion [22–24]. This may represent a compensatory mechanism to limit immune-mediated tissue damage. Our comparison to proteomic data from the TULIP-1 trial showed that LAG3 was downregulated by anifrolumab. In addition to an interferon-driven ISP, we also directly observed upregulation of interferon proteins themselves. Notably, IFNL1, a type III interferon, was strongly upregulated in both comparison to HVs and the analysis of disease activity. IFNL has received less attention than type 1 interferons in lupus pathogenesis, but its upregulation in SLE has been previously reported [25–29].

Our SomaScan results recapitulated historical studies of the complement pathway proteins beyond intact C3 and C4 (which are currently used in clinical practice as biomarkers) [30]. We observed positive association of C3d and negative association of CFP (properdin) with active disease, in keeping with previous reports [31–34]. C1Qc was one of the most strongly associated proteins with active disease. A previous study reported that C1Q negatively regulates type 1 IFN production by promoting preferential binding of immune complexes (ICs) to monocytes and thus reducing IC activation of pDCs, which are a major producer of type 1 IFNs [35]. However, we observed a strong positive correlation between C1Qc and levels of ISPs. Our data are consistent with the observation of reductions in whole-blood *C1Qc* gene expression following anifrolumab therapy [20].

Disease activity-associated proteins included several members of the TNF superfamily cytokines and receptors. One example was TNFSF4 (OX40L), whose plasma levels increased in more active disease. The *TNFSF4* gene locus is a susceptibility locus for SLE, pointing to its causal role in SLE aetiology [36]. In keeping with this, conditional knock out of B cell OX40L ameliorates mouse models of SLE [37]. Our data add to these lines of evidence suggesting that OX40L is a potential therapeutic target in SLE. Another example was TNFSF15 (TL1A), which is a promising therapeutic target in inflammatory bowel disease [38]. Upregulated TNFSF receptors included TNFRSF1B (TNF-R2), which acts a receptor for both TNF and lymphotoxin alpha (LTA), and TNFRSF10B, the receptor for the apoptosis-inducing cytokines TRAIL. Interleukins increased in active disease included IL6, IL11 and IL24. Conversely, there were negative correlations between disease activity and the anti-inflammatory proteins IL1R2 and IL10RA. IL1R2 is a decoy receptor for IL-1, and reduced IL1R2 would be expected to increase pro-inflammatory IL-1 signalling. IL10RA is a receptor for IL-10, an important anti-inflammatory cytokine. Genetic variants in the *IL10RA* gene are associated with early-onset inflammatory bowel disease [39]. Our findings raise the possibility that insufficient activation of homeostatic mechanisms contribute to increased disease activity, although since our data are observational, we cannot exclude the possibility that reductions in anti-inflammatory proteins are secondary to active disease rather than contributors to it.

Co-expression network analysis identified modules of correlated proteins. Two modules had notable associations with clinical and laboratory parameters. One module correlated with the ISG and multiple autoantibodies, consistent with previous reports linking high ISG expression to autoantibodies to Ro, U1-RNP, Sm, and dsDNA [40, 41]. Its hub protein, ISG15, bridges the type 1 and type 2 interferon pathways: induced by type I interferons, it provides negative feedback to downregulate type I responses while promoting interferon-γ [42]. A previous study identified ISG15-secreting plasmablasts and plasma cells in active SLE [43]. The second protein module reflected renal function and other renal-related physiology, and was enriched for ECM-and fibrosis-related pathways, suggesting it captures renal scarring.

Serological profiles in SLE correlate with clinical phenotype and prognosis [17, 18]. For example, anti-dsDNA antibodies are associated with renal disease and anti-Sm antibodies with poor prognosis. Here we show distinct proteomic signatures associated with anti-Sm and anti-dsDNA antibodies that persist after adjustment for the potential confounder of disease activity. Anti-Sm positive SLE showed increased ISPs and reduced circulating snRNPs, possibly reflecting depletion by anti-Sm antibodies targeting epitopes on small nuclear ribonucleoprotein complexes. Although the reductions in snRNPs were in plasma, this may proxy intracellular abundance, raising questions about anti-Sm effects on cellular machinery. Anti-dsDNA positive SLE was associated with elevation of the neutrophil protease, proteinase-3 (PRTN3). This might reflect release of neutrophil contents via neutrophil extracellular traps (NETs), which are a well-established source of nuclear autoantigens in SLE. It is also possible that anti-dsDNA antibodies drive further NET formation and release of proteinase-3.

CD40LG showed strong associations across multiple analyses, including correlations with disease activity, dsDNA antibody status, and complement C3/C4 levels. Intriguingly, the association with disease activity was specific to individuals with dsDNA antibodies. CD40LG/CD40 interaction drives B cell differentiation into immunoglobulin (Ig)G-secreting plasma cells, making it central to antibody responses. This is consistent with the associations with dsDNA antibodies (both positivity and titre) and reductions in complement C3/C4 (reflecting complement consumption by immune complexes). Previous studies have identified increased expression of CD40 or CD40LG in immune cells and serum in a range of immune-mediated diseases, including SLE [44], and, in line with our data, Goules et al found that CD40LG was elevated in the sera of SLE patients and associated with dsDNA positivity and disease activity [45]. Further support for a pathogenic role for the CD40 pathway in SLE comes from animal models; blocking the CD40-CD40LG interaction ameliorates lupus-like disease in NZB/SWR or NZB/NZW F1 mice [46, 47] and also reduced dsDNA antibodies. There have been efforts to therapeutically target CD40LG in SLE and other autoimmune diseases. While first generation anti-CD40LG monoclonal antibodies (mAbs) were abandoned due to thrombotic events [48], direct targeting of CD40 or newer generation anti-CD40LG mAbs bioengineered with inert or absent Fc tails circumvents this issue. A phase 2 trial of the anti-CD40 mAb BI655064 in lupus nephritis failed to meet its primary end-point, although post-hoc analysis suggested potential benefits of higher doses [49]. Evaluation of Frexalimab, a bioengineered anti-CD40LG mAb is ongoing (NCT05039840). Notably, comparison of our data to proteomic data from the TULIP-1 study revealed that, unlike many of the proteins we found associated with disease activity, CD40LG was unaffected by anifrolumab. This suggests that CD40LG axis inhibition might be a complementary approach to anti-IFN therapy by tackling an orthogonal pathogenic pathway. Given the strong associations between CD40LG and dsDNA Ab and complement levels, our data highlight that a stratified medicine approach for anti-CD40 pathway therapeutics might be beneficial. This concept has parallels to the path to approval of belimumab, which proved effective in SLE when used in serologically defined subsets of patients after initial trial failure [50].

Our study had several limitations. Although our data were split into a discovery and replication subcohorts for case versus control analysis, this was a single-centre study. For within-cases analyses, we analysed both subcohorts together to boost power and thus we did not have a hold-out set for validation. Patients were sampled opportunistically at various points in the disease course and not at pre-defined timepoints; while this enabled cross-sectional evaluation of markers of disease activity, disease duration, chronic damage and past and current treatment were potential confounders. We did not have sufficient longitudinal samples to formally and systematically evaluate intra-individual changes in disease activity-related markers over time. Future studies using inception cohorts with serial sampling are needed to validate putative biomarkers of disease activity identified in the present study. Finally, interpretation of plasma proteomic measurements can be challenging. In addition to canonical soluble proteins, the SomaScan platform includes targets measuring intracellular and membrane-bound proteins. Their detection may represent release from dead cells. Moreover, where proteins exist in both membrane-bound and cleaved states, it is not always clear whether plasma proteomic assays are exclusively capturing the soluble form or also protein from cell membranes (for example, arising from in vivo sources such as exosomes or from sample handling). Complementary cytometry data on paired samples would provide additional insights.

In summary, this study provides the most comprehensive map of the plasma proteome in SLE to date. Stratification by autoantibody status reveals distinct proteomic endotypes even after adjustment for disease activity. Our data provide insights into pathogenesis and highlight potential therapeutic targets.

## Methods

### Subjects and Samples

We recruited 268 patients with SLE from the Imperial Lupus Centre at the Hammersmith Hospital, London, United Kingdom (UK). All patients were anti-nuclear antibody positive and fulfilled the 1997 American College of Rheumatology criteria. We recruited 86 healthy volunteers (HVs) to provide a control group, sampled as two temporally distinct groups comprising 45 and 41 individuals, respectively. No patients had received anifrolumab since this drug is not licensed in the UK.

Human samples used in this research project were obtained from the Imperial College Healthcare Tissue and Biobank (ICHTB). ICHTB is supported by the National Institute for Health Research Biomedical Research Centre based at Imperial College Healthcare NHS Trust and Imperial College London. ICHTB is approved by Wales REC3 to release human material for research (22/WA/0214). Ethical approval was provided by the ICHTB (Human Tissue Authority Licensing number 12275; Project number R14042-3A; sub-collection IMM_MB_13_001) and informed consent obtained from all participants. Peripheral blood was collected in EDTA tubes and centrifuged at 1,000G for 10 minutes at room temperature within three hours of venepuncture and stored at −70°C without freeze-thaw before assay processing.

### Proteomic measurements and quality assessment

Plasma proteins were measured using the aptamer-based SomaScan v4.1 platform (Boulder, Colorado, USA). The SomaScan v4.1 platform uses modified aptamers (SOMAmers) that bind to their targeting proteins for measurement. A total of 7,288 SOMAmers targeting 6,595 unique human proteins were measured (some proteins were assayed by multiple distinct SOMAmers; **Supplementary Data 15**). To avoid interference of anti-dsDNA autoantibodies with the SOMAmers, a validated anti-DNA antibody protocol was used [10]. This involved adding herring sperm DNA to saturate anti-dsDNA autoantibodies within each sample and prevent these antibodies from binding the SOMAmers. The platform contains buffer replicates containing no protein which are used to estimate the background noise level, calibrator replicates to control for intra-plate variations and batch effects across experiments, and quality control (QC) replicates for signal normalisation and QC check. Multiple normalisation and calibration steps were performed using SomaLogic’s standard protocol. Data normalised by adaptive normalisation by maximum likelihood was used for our analyses. Relative fluorescence units were log_2_-transformed before analyses. Proteomic measurements were performed in two separate batches: batch A consisted of 226 SLE and 45 healthy samples and batch B of 57 SLE and 41 HV samples.

In addition to the manufacturer’s quality assessment, we performed additional steps including PCA and inspection of sample distributions via boxplots. We identified batch effects, with separation of Batch A and Batch B on PCA. **(Supplementary Figures 12A-B)**. After manual examination on the PCA plot, three samples from Batch A were removed since they were distinctly distributed from the main cohort of samples (>3 standard deviations from the means of PC1 and PC2) and also processed on the same day, suggesting technical rather than biological variation (**Supplementary Figure 12A**). Additionally, 1 sample from batch A and 4 samples from batch B were removed since they were flagged as potential poor quality samples by SomaScan’s standard QC pipeline requiring extremely high (>2.5) or low (<0.4) scaling factors for normalisation. For 3 patients, serial longitudinal samples were available. In these instances, we retained the first sample for each patient and excluded the others from all downstream analyses except for the evaluation of intra-individual changes over time in disease activity-associated markers (see below). For the main analyses (excluding serial samples), post QC samples sizes were as follows: Batch A: 207 SLE, 45 HVs and Batch B: 53 SLE, 41 HVs.

### Protein nomenclature

Due to non-standardisation of protein nomenclature (e.g. where a given protein has multiple names) and the lack of human-interpretability of Uniprot identifiers, we annotated each protein using the non-italicised gene HUGO symbol of the encoding gene. In instances where there were multiple distinct SOMAmers mapping to the same HUGO gene symbol (1,669 SOMAmers mapped to 781 unique gene symbols and 782 Uniprot identifiers), we concatenated the gene symbol with the SOMAmer identifier to preserve a unique identifier for each analyte. Importantly, in some instances, multiple SOMAmers mapping to the same gene symbol represented measurements of distinct biological entities. For example, the intact complement C3 protein and its distinct cleavage products such as C3a, C3b, and C3d, all mapped to a single HUGO gene symbol and Uniprot ID. However, in other instances there were multiple SOMAmers that are nominally directed against the same target protein. A comprehensive annotation file mapping of SOMAmers to their target proteins, Uniprot IDs and corresponding gene IDs, and gene symbols is provided in **Supplementary Data 15**.

### Complement proteins

Various complement proteins and cleavage fragments were included on the SomaScan platform. To evaluate the accuracy of complement C3 and C4 quantification, we compared the levels of intact complement C3 and C4 proteins measured by SomaScan in SLE patients to their corresponding values measured in the clinical laboratory from the same blood draw. We performed this analysis separately for each batch to avoid variation related to batch.

### Identification of differentially abundant proteins in SLE vs healthy samples

For each protein measured, multiple linear regression was used to analyse differences in protein abundance between SLE and HVs samples using the lm() function in R. Age and sex were adjusted for by including them as covariates. The regression model in Wilkinson notation was:

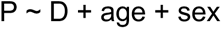

where P was plasma level for a given protein and D was a binary variable disease status (SLE vs healthy).

In Batch B, we observed inflation of test statistics and an unusually high proportion of proteins that were significantly differentially abundant in the comparison of SLE vs HVs (**Supplementary Figures 13-14**). We also observed marked separation of SLE and HV samples on PC1 (when PCA was performed limiting to Batch B; **Supplementary Figure 15**), with PC1 accounting for 42% of the variance. The most upregulated protein was PGAM1, which has been identified by Somalogic as a marker of pre-analytical variation including time to spin, suggesting that systematic non-biological variation was confounding the differences between SLE and HVs in Batch B (**Supplementary Figure 13)**. We therefore included the first principal component as a covariate to adjust for pre-analytical variation in Batch B (see **Supplementary Note**).

### Statistical significance

P-values were adjusted using the Benjamini-Hochberg (BH) method (P_BH_) to control the false discovery rate (FDR) across multiple tests. We defined statistical significance as P_BH_ <0.05 (i.e. leading to an FDR <0.05) for all analyses in this study.

### Identification of proteins associated with disease activity

Disease activity was assessed using the SLE Disease Activity Index (SLEDAI)-2K [51]. Given that SLEDAI-2K scores have a highly non-normal distribution and can be sensitive to the weightings attributed to specific clinical features, prior to testing for association with proteins, we categorised disease activity into a four level ordinal variable as follows: ‘high disease activity’ (HDA) was defined as SLEDAI-2K ≥ 10; ‘moderate disease activity’ (MDA) as SLEDAI-2K between 5 and 9; ‘low disease activity’ (LDA) as SLEDAI-2K ≤ 4 but not meeting the definition for remission; ‘remission’ as defined by the Definition of Remission in SLE (DORIS) criteria [52], characterized by clinical components of SLEDAI-2K = 0 (i.e. not including dsDNA antibody or complement status) and a prednisolone dose ≤ 5 mg/day.

To identify proteins associated with disease activity, we performed multiple linear regression for each protein using the lm() function in R. Protein abundance was regressed on disease activity (coded as an ordinal variable: “In remission”, “LDA”, “MDA”, “HDA”), with sex and batch as covariates. As a sensitivity analysis, Spearman’s rank correlation was conducted between protein abundance and SLEDAI-2K scores. Prior to correlation testing, protein abundances were adjusted to account for sex and batch effects using a linear model and the resulting residuals were used in the Spearman’s correlation test versus SLEDAI-2K scores.

### Exploratory Longitudinal analysis

A subset of disease activity–associated proteins was selected for further investigation in a longitudinal analysis of patients (n=3) with samples taken at four or more timepoints. We first selected proteins that were: (i) potential drug targets, defined as having an associated compound with a characterized mechanism of action in Open Targets (Dataset: “Drug - mechanism of action”; data version 25.03), and (ii) were members of the pathways significantly enriched among disease activity–associated proteins. From this list we then selected the 5 proteins with the greatest effect sizes in the ordinal linear regression analysis testing for association with disease activity. These criteria resulted in 5 proteins, one of which was an interferon-stimulated protein (CXCL10). In addition, we added two interferon-stimulated proteins that were significantly associated with disease activity. Repeated measures correlation was used to assess the association between these proteins and clinically measured disease activity traits, accounting for the non-independence of observations due to repeated measurements within the same individuals [53]. This approach is more appropriate than Pearson’s correlation, which assumes independent observations.

### Identification of proteins associated with autoantibody status

We analysed proteomic associations with the presence of antibodies to Smith (Sm), anti-dsDNA and phospholipids. For each plasma protein, abundance (P) was regressed on antibody status (Ab; a binary variable) with batch and sex included as covariates. The regression model in Wilkinson notation was P ∼ Ab + batch + sex. To identify associations with Sm antibodies independent of disease activity, we then repeated the analysis adding disease activity status as a covariate to the model.

For the dsDNA analysis, samples were classified as anti-dsDNA negative if the level was lower than 10 IUperML (the manufacturer’s defined threshold) and as anti-dsDNA positive if the patient’s plasma anti-dsDNA autoantibodies level was higher than 20 IUperML. Patients with anti-dsDNA levels (n=40) within the borderline range of 10-20 IUperML were excluded in this analysis. To identify proteins associated with anti-dsDNA positivity independent of disease activity, we used a modified SLEDAI-2K score excluding the anti-dsDNA component from the score to avoid over-adjustment since SLEDAI-2K includes 2 points for dsDNA antibody positivity. We re-calibrated the disease activity category thresholds between LDA, MDA and HDA based on this modified score (DORIS remission definition was unchanged since this does not include anti-dsDNA status). Specifically we normalised the modified SLEDAI-2K by the highest possible score excluding dsDNA antibody positivity (i.e., 105-2 = 103), and applied the same procedure to the unmodified SLEDAI-2K thresholds used to define activity categories described earlier. To illustrate this procedure, an individual positive for anti-dsDNA antibodies with an unmodified SLEDAI-2K score of 11 would have been defined as being in HDA (defined as SLEDAI-2K of 10 or higher) in the earlier analyses. The modified SLEDAI-2K score would be 9. The normalised score would be 9/103=0.087. The new cut-off for HDA would be 10/103=0.097. Thus this individual would be classified as in MDA for the purposes of covariate adjustment in the analysis testing for proteins associated with anti-dsDNA antibodies independent of disease activity.

As a sensitivity analysis, we tested protein abundances against anti-dsDNA titre (as a continuous variable) using all available SLE samples. To do this, protein abundances were first adjusted to account for sex, batch and disease activity (as an ordinal variable) using a linear model and the resulting residuals were used in the Spearman’s correlation test versus anti-dsDNA titre.

To formally test whether CD40LG’s association with disease activity was dependent on dsDNA antibody status, we fitted a regression model with an dsDNA antibody status as a binary variable (Ab) x disease activity interaction term:

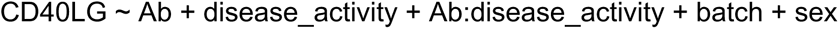

A significant interaction term indicates that the effect of disease activity on the protein is dependent on dsDNA antibody status.

### Pathway enrichment analyses

For SLE-vs-HVs comparisons and disease activity analysis, we performed Gene Set Enrichment Analysis (GSEA) using the fgsea R package (version 1.24.0) [54] and the Human MSigDB Collection “C2: curated gene sets” (sub-collection: “Canonical pathways”) which consists of 4,023 gene sets (v2025.1) mainly sourced and curated from BioCarta, Kyoto Encyclopedia of Genes and Genomes, Pathway Interaction Database, Reactome, and WikiPathways [55]. For SLE-vs-HVs comparisons, GSEA was performed separately in each batch. We then used robust-rank aggregation (RRA) [56] to identify the pathways most consistently enriched across both batches. RRA outputs a score analogous to a P-value for each pathway term. These scores were −log10 transformed to represent the aggregated score. A higher aggregated score indicates stronger consistency.

Over-representation tests were used to identify enriched pathways in WGCNA protein modules and antibody-associated protein lists, implemented using the ‘enrichPathway’ function from the ReactomePA R package (v1.42.0) [57]. Proteins were first mapped to corresponding Entrez IDs since this package requires Entrez ID annotation. Entrez IDs mapped to proteins in the module were tested against the background set of 6,363 Entrez IDs representing the proteins measured on SomaScan v4.1. All pathway enrichment analyses were restricted to protein sets containing 10–500 proteins to exclude very small sets vulnerable to noise and large sets representing broad and non-specific processes.

### Protein network analysis

We applied the Weighted Gene Co-expression Network Analysis (WGCNA) package (v1.72-5) [58] to identify modules of correlated proteins. For this analysis we used SLE samples (n=207) from Batch A. A soft thresholding power of 6 was selected based on the criterion of scale-free topology and mean connectivity. Networks were constructed and modules were identified by the adjacency and topology overlap matrices, with minimum module size set to 20. We then refined the modules by their similarity based on modular eigenprotein (representing the first principal component of each module’s protein abundance profile) correlation using a dynamic tree-cut approach. Modules were merged if their eigenprotein-based dissimilarity was below a threshold of 0.25. The resulting modular eigenproteins were then used to correlate with 20 clinical traits using Spearman’s rank correlation test, including 11 continuous traits: age, SLEDAI-2K, neutrophil count, lymphocyte count, haemoglobin, creatinine, urine protein-creatinine ratio (uPCR), albumin, complement C3, complement C4, and whole-blood ISG score (see below), and 9 binary traits: positivity of antibodies against cardiolipin (ACA), RNP-A, RNP-68, Sm, La, Ro52, Ro60, dsDNA, and sex. Correlations with P_BH_ < 0.05 were considered significant. Eigenprotein-based connectivity, or module membership, was calculated for each protein in each module by correlation between the protein abundance levels and the corresponding module’s eigenprotein values. The protein with highest module membership was then defined as the hub protein for each module. The network of the proteins in the selected module was constructed using the topology overlap matrix which calculates the strength of similarity between each pair of proteins. The network was then visualised in Cytoscape (v3.10.3) [59], with nodes representing proteins and thickness of edges representing the strength of similarity. Only the edges with a threshold value of > 0.03 were included in the visualised network.

### Whole-blood interferon stimulated gene score

The whole-blood interferon-stimulated gene score was measured as described previously [60]. In brief, total RNA was isolated from whole blood samples using Tempus RNA isolation kits (Thermo Fisher Scientific) following the manufacturer’s protocol. Gene expression was measured by real-time qPCR and relative gene expression of *CMPK2*, *EPSTI1* and *HERC5* were calculated by subtracting the housekeeping gene *HPRT* using the ΔCt method. The ISG score was calculated using the mean ΔCt from *CMPK2*, *EPSTI1* and *HERC5* genes *(−1).

## Data Availability

Summary-level data are contained in the supplementary data.

## Resource availability

### Lead contact

Requests for further information and resources should be directed to the lead contact, James E. Peters.

### Materials availability

This study did not generate new unique reagents.

## Acknowledgements

The study was funded by a Medical Research Foundation Fellowship to J.E.P. (MRF-057-0003-RG-PETE-C0799) and by the NIHR Imperial Biomedical Research Centre Immunology Theme. The views expressed are those of the authors and not necessarily those of the NIHR or the Department of Health and Social Care. M.C.P. is a Wellcome Trust Senior Fellow in Clinical Science (grant no. 212252/Z/18/Z). We thank Shanice Lewis for technical assistance.

