## Supplementary Material for "Mapping the plasma proteomic architecture of systemic lupus erythematosus"

### Supplementary material for ‘Mapping the plasma proteomic architecture of systemic lupus erythematosus’ by Leung et al

Table of Contents

**Supplementary Note**

**Supplementary Figures**

**Titles for Supplementary Data**

#### Supplementary Note

Correction for pre-analytical variation confounding SLE vs HV differential abundance analysis in Batch B

Principal component analysis performed separately on each batch of data showed that the SLE were sharply demarcated from HV samples in Batch B on plots of PC1 vs PC2. This pattern was not observed in Batch A, where SLE and HV samples were more overlapping on the PCA plot (**Supplementary Figure 15**). Moreover, PC1 explained 42% of the variance in the dataset in Batch B.

Differential protein abundance testing of SLE vs HVs in Batch B resulted in a very high proportion of significantly differentially abundant proteins (DAPs). 5,643 (77%) of 7,288 measured proteins) were significantly differentially abundant at 5% false discovery rate (**Supplementary Figure 13**). This resulted in an unusual appearance of the volcano plot. In contrast, in Batch A, only 1,821 proteins were significant, despite a much larger sample size (207 SLE, 45 HVs) and hence statistical power. We confirmed the impression of inflated signals in Batch B using QQ plots (**Supplementary Figure 14**). This finding could not be explained by differences in disease activity (median SLEDAI = 4 in batch A, median SLEDAI = 2 in batch B; P = 0.07) or other clinical characteristics between the Batches. We performed a number of sensitivity analyses, including restricting to females only, but the inflated signals in Batch B persisted. For SLE samples, we had contemporaneous clinical laboratory measurements of complement C4 available and so tested these with SomaScan-measured complement C4. This revealed very strong correlations between SomaScan and clinical lab measurements of C4 in both Batch A and Batch B, suggesting that the SomaScan measurements were reliable in the SLE samples (**Supplementary Figure 3B**). This led us to suspect that the source of inflation might be the pre-analytical variation related to the HV samples in Batch B. Potential sources of variability for HV samples in Batch B included differences in personnel performing sample processing and time to spin. In contrast, for Batch A HV samples and SLE samples the sample handling was more consistent. Moreover, PGAM1 was most upregulated with the greatest effect estimate ($\log_2$ fold change) in SLE vs HVs in batch B samples. This marker has been identified by SomaLogic as suggesting pre-analytical variation and is affected by time to spin.

Given this and the separation on the PCA plots, we computed the first principal component (PC) for all batch B samples and included it as a covariate in linear regression models for SLE vs HV differential abundance testing in the batch B analysis. Following PC-adjustment, the proportion of significantly differentially abundant proteins at 5% FDR fell to 5.9% and the QQ plots were improved **(Supplementary Figures 13-14)**. In addition, the most strong signals were interferon-stimulated proteins, i.e. a biological plausible finding.

Comparison of beta estimates and significant proteins (5% FDR) between the two batches are shown in **Supplementary Figure 13**. Without PC1 adjustment in Batch B, 4,182 proteins were significant in Batch B but not Batch A, and many proteins with large effect sizes in Batch B had no significant effect in Batch A. Following PC1-adjustment in Batch B, there was greater concordance of effect size estimates.

Exploratory Longitudinal Analysis

For 3 patients, longitudinal samples taken at 4 or more timepoints were available. The limited sample size precluded formal biomarker analysis, but we used these data to qualitatively explore selected proteins that we had identified as disease activity-associated in the cross-sectional analysis. Five proteins that are potential drug targets, involved in the enriched pathways, and with largest effect sizes were selected, comprising CXCL10, CCL2, CD40LG, HAVCR2, and C3d (measured by SOMAmer seq.5803.24). Two disease activity-associated ISPs (MX1 and B2M) were also included. We visualised the longitudinal relationships between these 7 proteins and SLEDAI scores and disease activity-related serological markers (clinically measured C3, C4, and anti-dsDNA autoantibody levels). Our data showed that the levels of these proteins generally tracked with changes in disease activity over time **(Supplementary Figure 7A)**. For example, in Patient 1, who exhibited a progressive decline in disease activity leading to remission, most of the selected proteins showed a consistent decrease in abundance across timepoints. Statistical evaluation with repeated measures correlation (accounting for non-independence of serial samples) [1] confirmed strong within-patient relationships: all proteins were significantly correlated with at least 1 clinical parameter, showing positive correlations with SLEDAI-2K and anti-dsDNA titres and negative correlations with C3 and C4 levels **(Supplementary Figure 7B)**. For example, CD40LG was strongly correlated with anti-dsDNA levels (r 0.95, $P_{BH}$ $5.6\times10^{-4}$; **Supplementary Figure 7C**) consistent with the key role of the CD40LG-CD40 pathway in B cell proliferation/differentiation and antibody production [2]. C3d was inversely correlated with clinically measured C3 (r -0.88, $P_{BH}$ $4.5\times10^{-3}$; **Supplementary Figure 7D**), in keeping with the known biology of the complement pathway (i.e. cleavage of C3 reduces the amount of intact C3 and increases C3d). Notably, several proteins returned to “normal” levels (within the IQR of HVs samples) as disease activity diminished. However, although levels of ISPs did broadly mirror disease activity within an individual, they largely remained elevated beyond the normal range.

#### Supplementary Figures

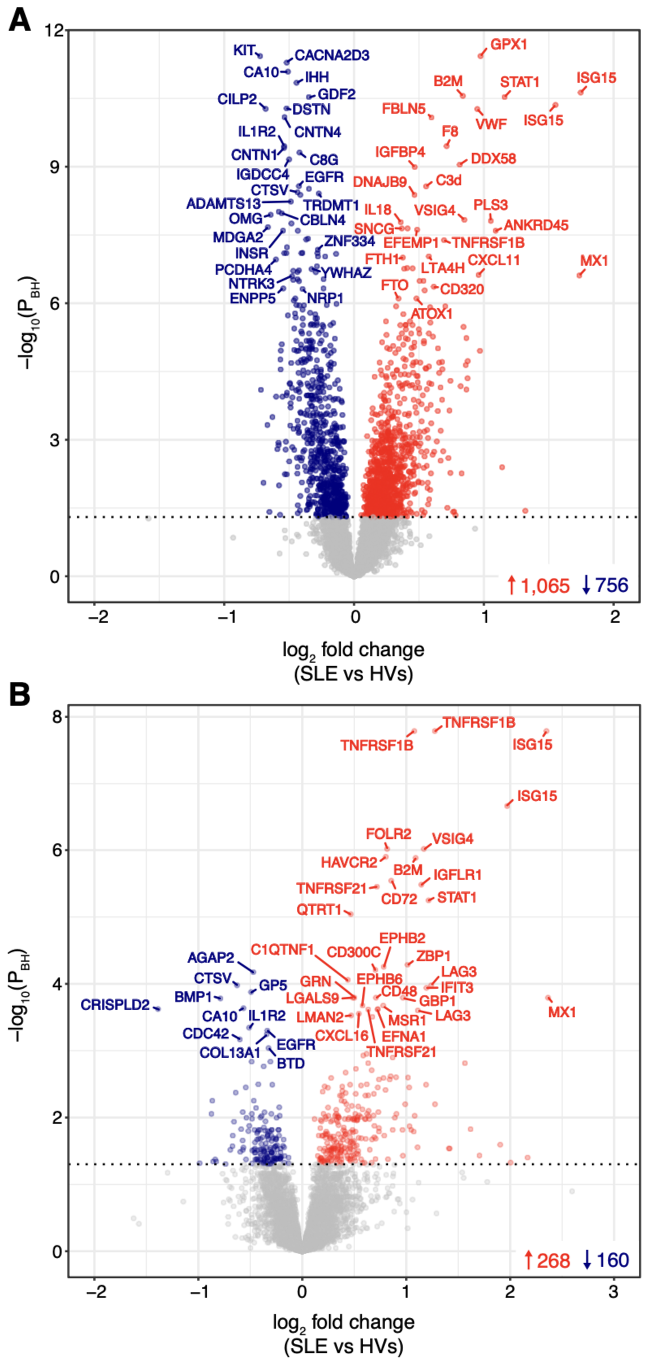


**Supplementary Figure 1. Differential abundance analysis of SLE vs HVs. A)** Volcano plot showing the $\log_2$ fold change and BH-adjusted P values of each protein from linear regression analyses in batch A. Age and sex were adjusted as covariates. Each point represents a protein. Significantly upregulated proteins are coloured in red while downregulated proteins are coloured in blue. Proteins with BH-adjusted P ≥ 0.05 are coloured in grey. **B)** Volcano plot showing the same information as in A, but the analysis was performed in batch B.

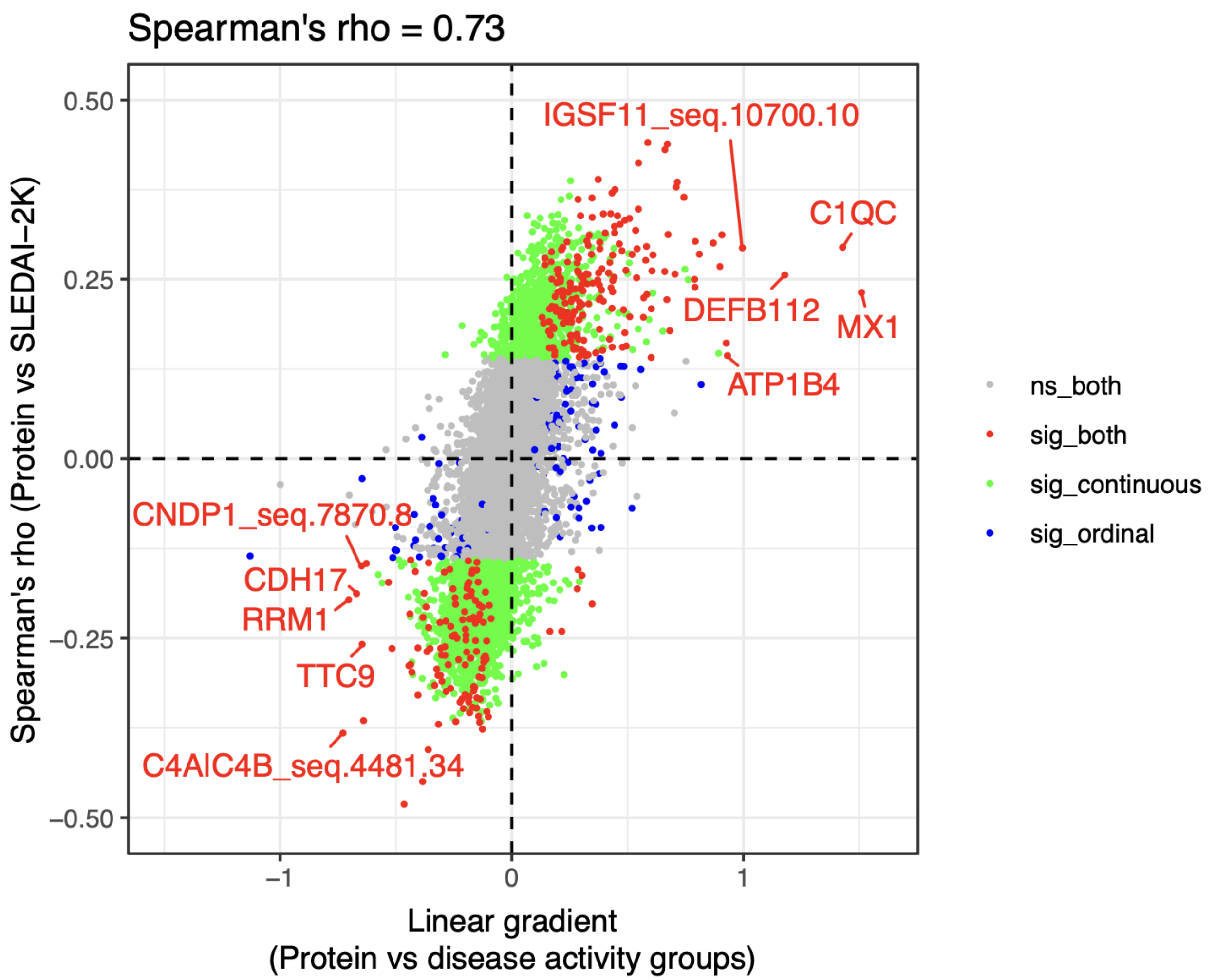


**Supplementary Figure 2. Sensitivity analysis treating SLEDAI as a continuous variable.** Sensitivity analysis comparing analysing disease activity as a 4-level ordinal variable versus using SLEDAI score as a continuous trait. X-axis: linear gradient obtained from linear regression model treating disease activity as ordinal variable. Y-axis: Spearman's correlation test on protein levels (residuals after adjustment for covariates) vs SLEDAI (as continuous trait). Each point represents a protein analyte and is coloured based on statistical significance - i) not significant (ns, FDR < 0.05) in both models (grey); ii) significant in both models (red); iii) significant only in the Spearman's test (green); or iv) significant only in the linear regression model (blue).

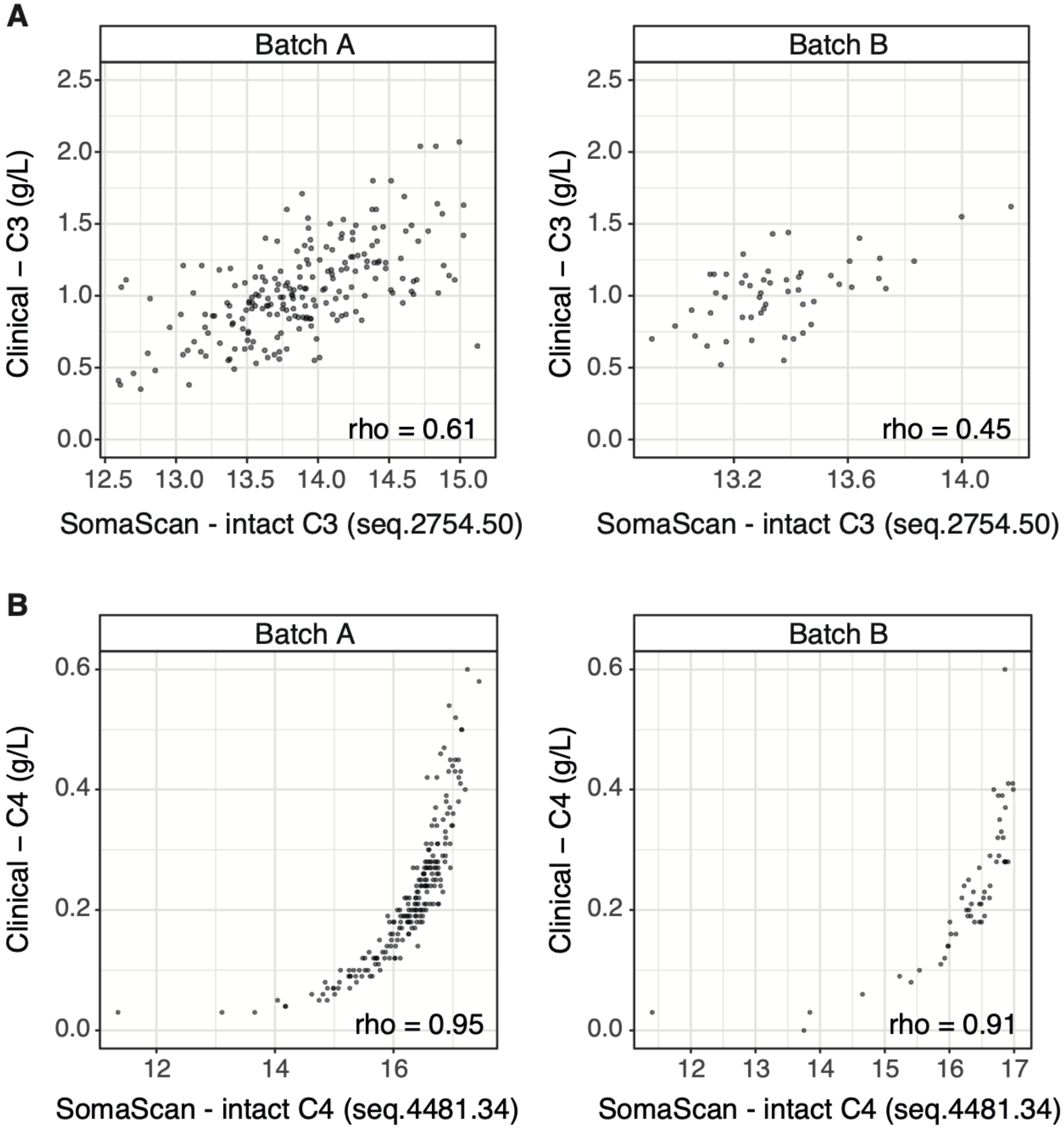


**Supplementary Figure 3. Correlation of SomaScan- and clinically measured complement levels. A)** Correlations between clinically measured C3 levels and SomaScan-measured intact C3 levels (measured by SOMAmer seq.2754.50) in both batches of data. **B)** Correlations between clinically measured C4 levels and SomaScan-measured intact C4 levels (measured by SOMAmer seq.4481.34) in both batches of data.

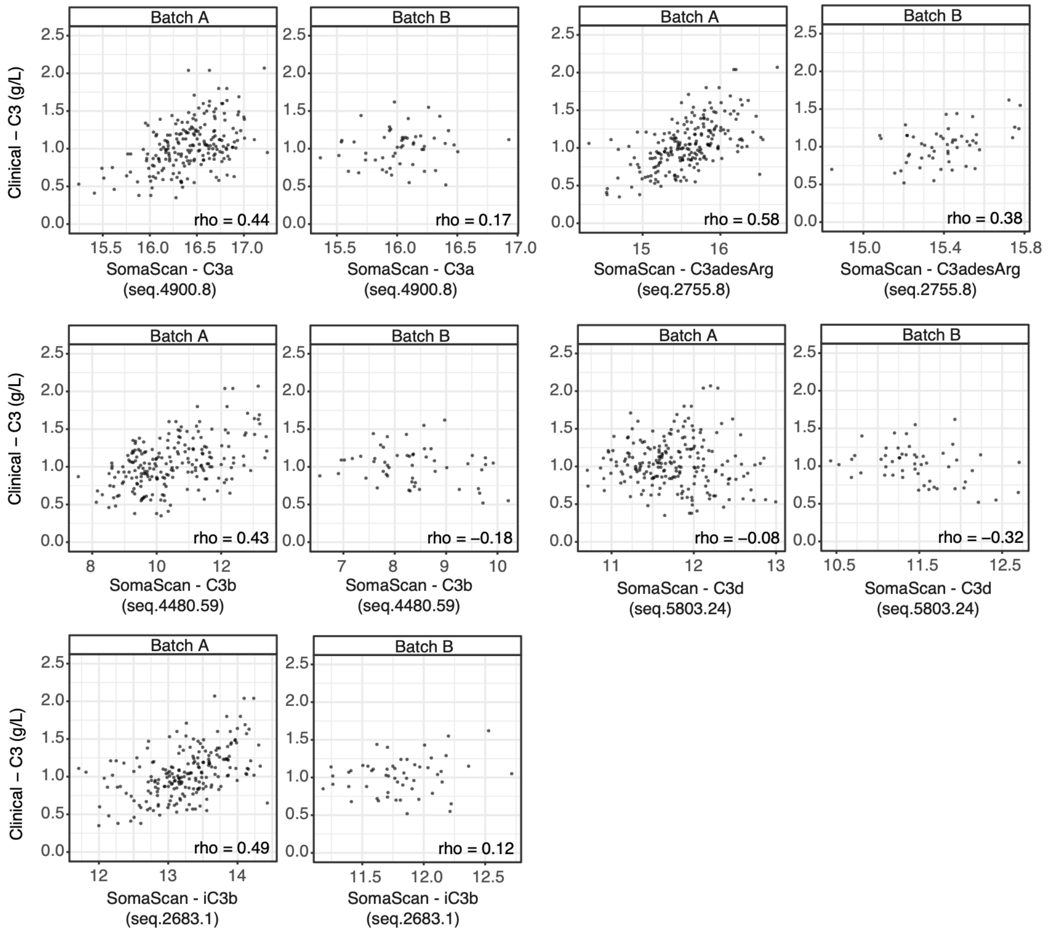


**Supplementary Figure 4. Complement cleavage fragments of C3 measured in SomaScan and their correlations with clinically measured intact C3 in two batches of data.** Five SOMAmers for C3 fragments were measured in SomaScan, targeting C3a (seq.4900.8), C3adesArg (seq.2755.8), C3b (seq.4480.59), C3d (seq.5803.24), and iC3b (seq.2683.1).

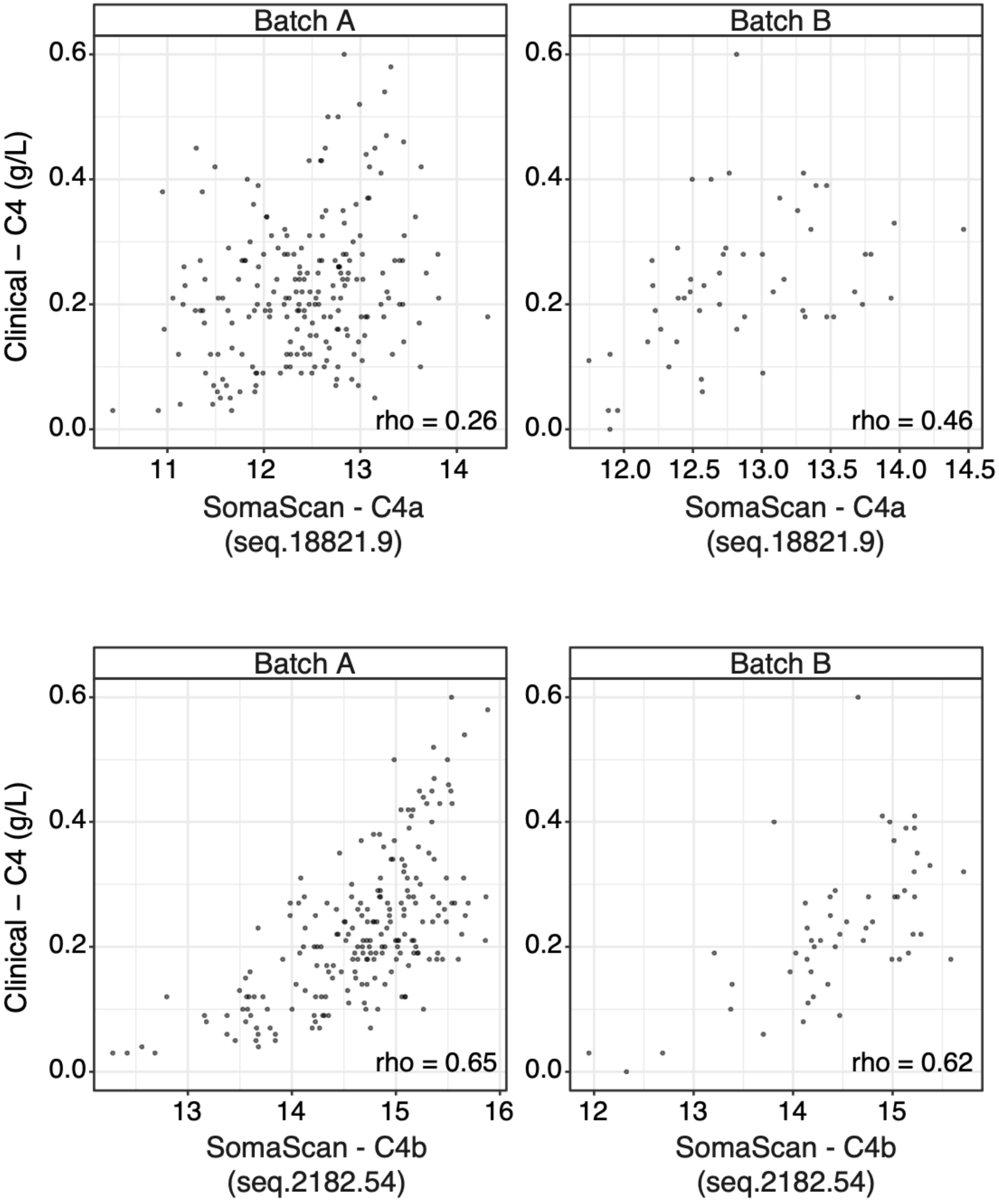


**Supplementary Figure 5. Complement cleavage fragments of C4 measured in SomaScan and their correlations with clinically measured C4 in two batches of data.** Two SOMAmers for C4 fragments were measured in SomaScan, targeting C4a (seq.18821.9), and C4b (seq.2182.54).

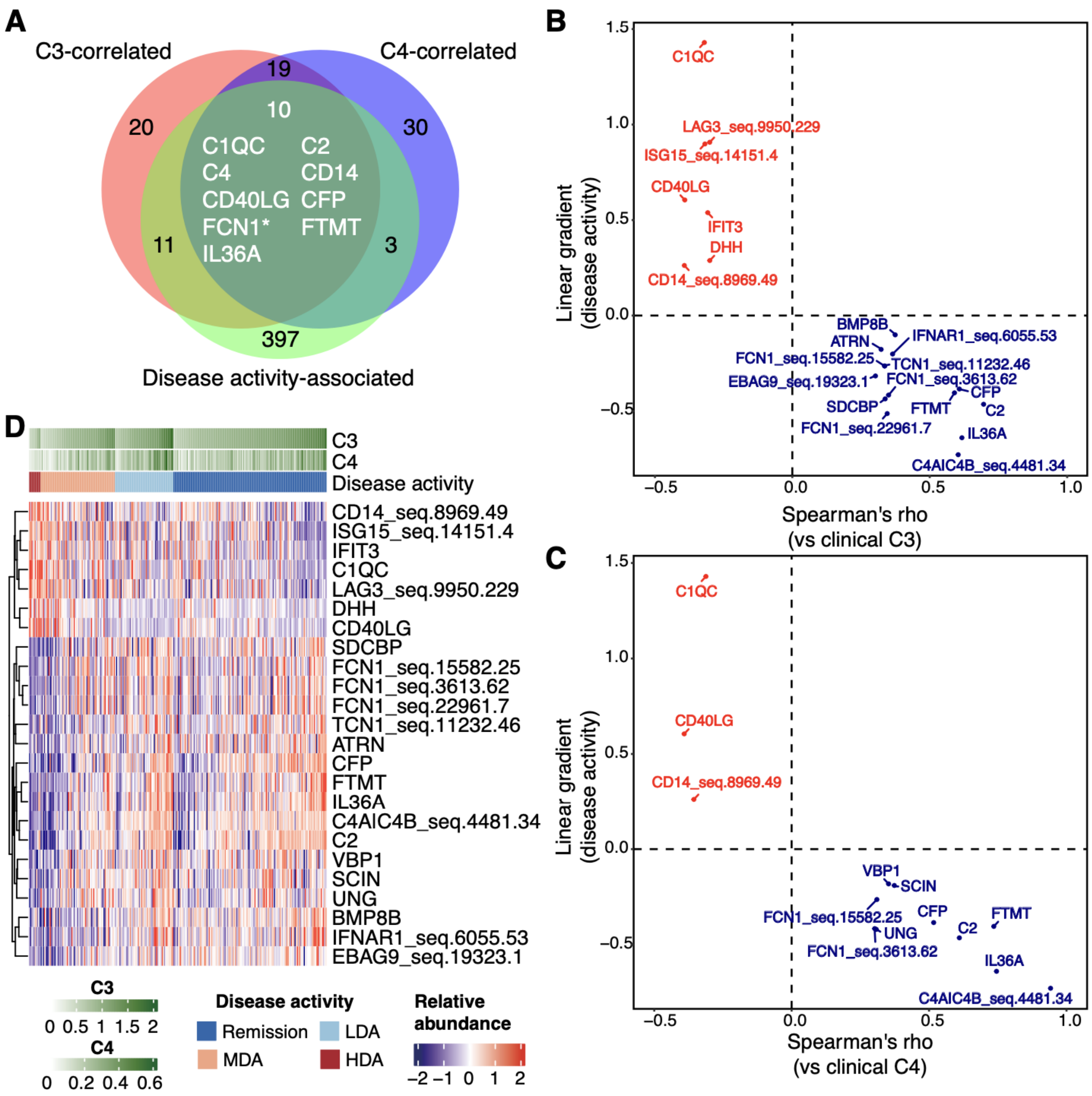


**Supplementary Figure 6. Proteins associated with disease activity and clinically measured C3 or C4. A)** Venn diagrams of the number of protein analytes significantly correlated with clinical C3 and C4 levels, and associated with disease activity. Ten SOMAmers correlated with both C3 and C4 and associated with activity were listed. *These include two SOMAmers for FCN1 (seq.15582.25 and seq.3613.62). **B)** Proteins associated with disease activity and with clinical lab-measured C3 levels. **C)** Proteins associated with disease activity and correlated with clinical lab-measured C4. Red: proteins positively associated with disease activity, Blue: proteins negatively associated with disease activity. **D)** Proteins associated with disease activity and with either clinical lab-measured C3 or C4. Protein levels were adjusted by sex and batch.

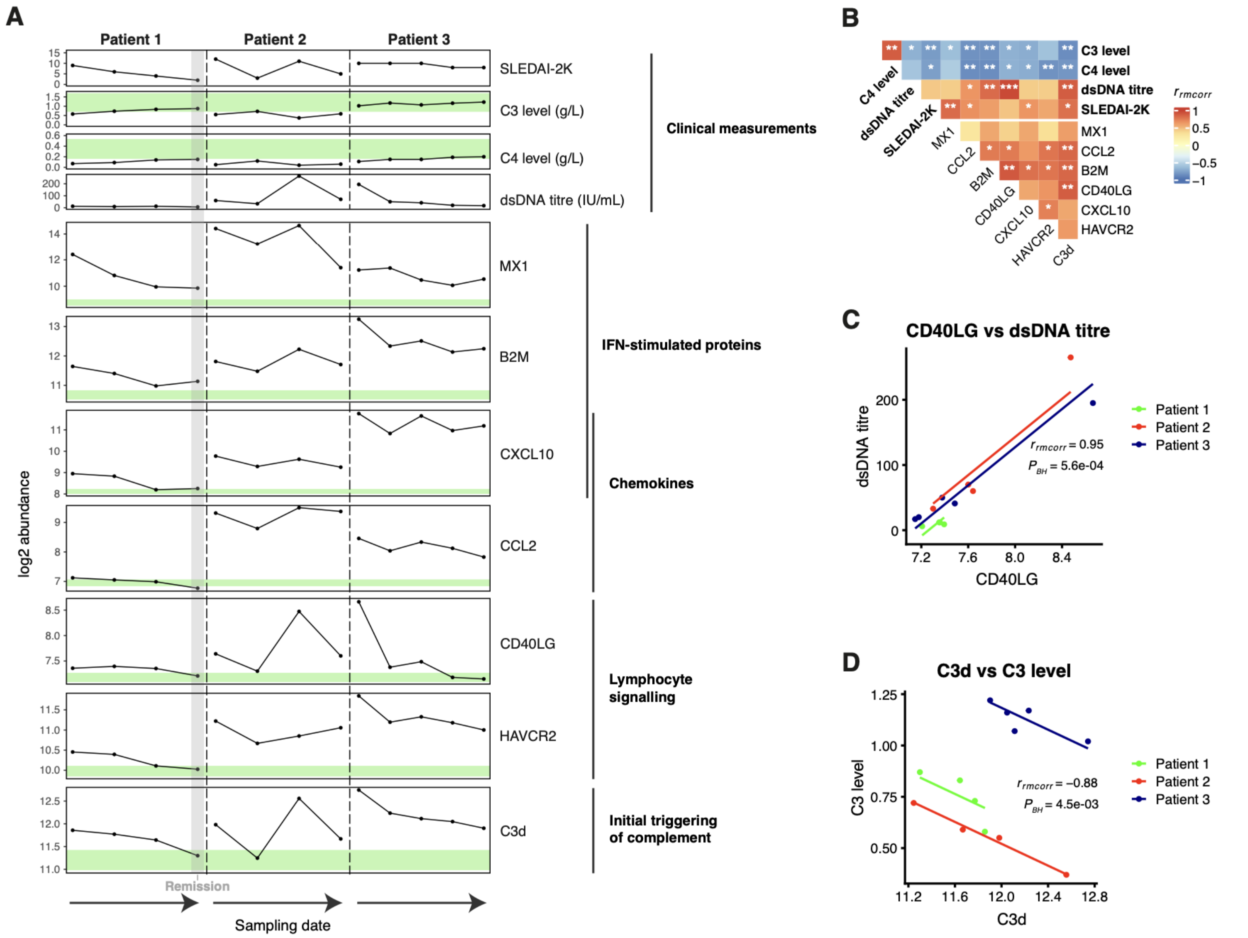


**Supplementary Figure 7. Disease activity-associated proteins in serial samples collected from 3 SLE patients. A)** Upper panel: SLEDAI-2K and clinically used disease activity biomarkers. Lower panel: plasma proteins measured using SOMAScan. Each point represents a sample. Samples were ordered by sampling date (from the oldest to the most recent one) within each patient. For clinical laboratory measured C3 and C4 levels, the green colour represents their normal reference range. For SOMAScan-measured plasma proteins, normal range was estimated using the median±interquartile range from the healthy volunteers. Proteins were grouped by their biological functions. The grey vertical shading area highlights the sample of Patient 1 taken during remission. Proteins B2M and HAVCR2 had multiple SOMAmers on the SOMAScan; in this figure B2M corresponds to measurement with SOMAmer “seq.3485.28” and HAVCR2 to “seq.5134.52”. **B)** Repeated measures correlations between clinical parameters (shown in bold font) and levels of SOMAScan-measured plasma proteins (non-bold) in all serial samples of the 3 patients. Red and blue represent positive and negative correlations, respectively. *, $P_{BH}<0.05$; **, $P_{BH}<0.01$; ***, $P_{BH}<0.001$. **C-D)** Intra-individual evaluation of longitudinal correlation between **C)** plasma CD40LG and anti-dsDNA autoantibody levels, and **D)** plasma C3d (SOMAScan) versus C3 levels (clinical laboratory measure).

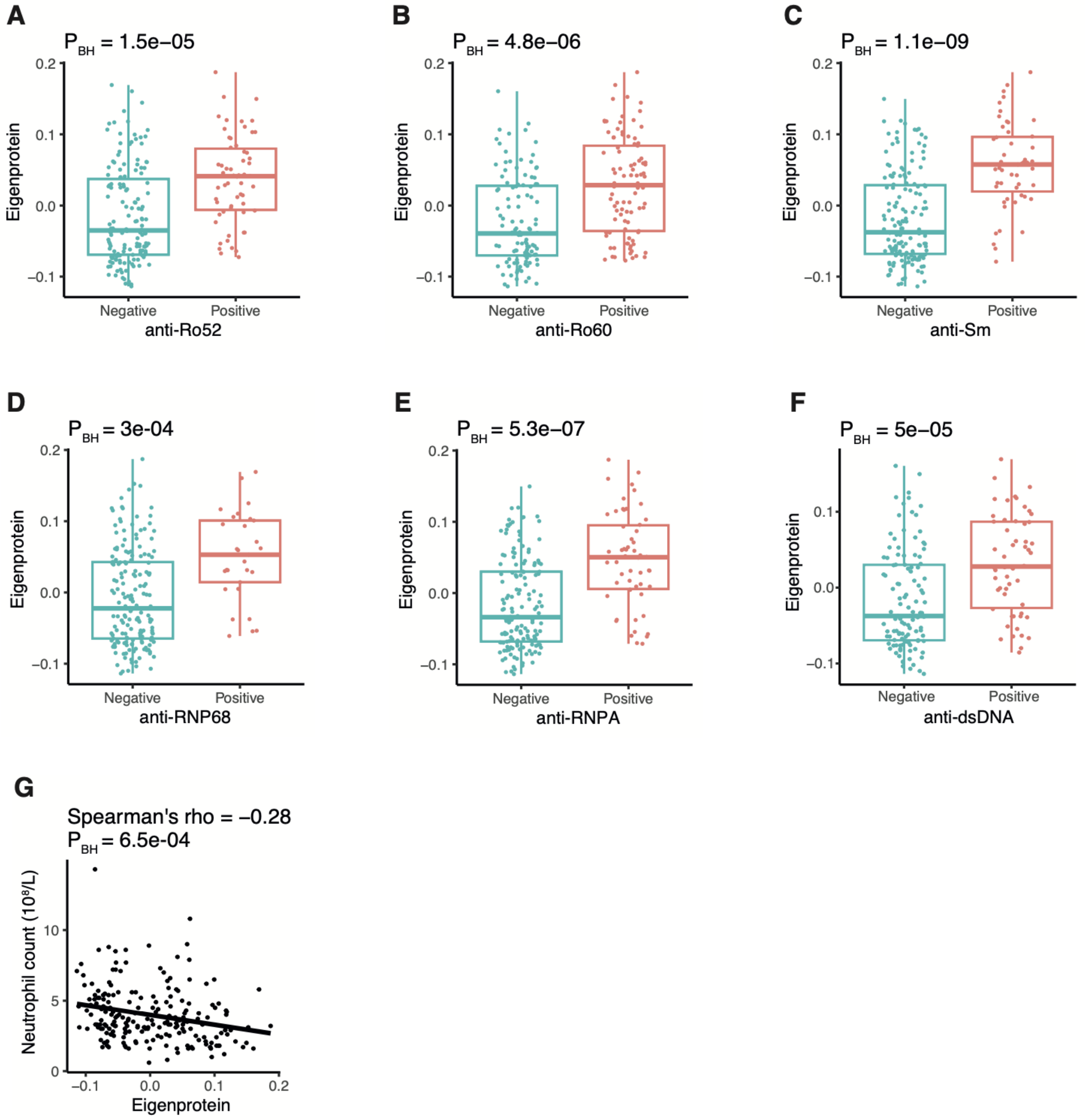


**Supplementary Figure 8. Clinical traits that were significantly associated with the red module identified by WGCNA.** All traits that were strongly associated (FDR < 0.001) with the modular eigenprotein values of the red module were shown, including 6 binary traits of autoantibodies status: **A)** anti-Ro52, **B)** anti-Ro60, **C)** anti-Sm, **D)** anti-RNP68, **E)** anti-RNPA, **F)** anti-dsDNA, and 1 continuous trait: **G)** neutrophil count.

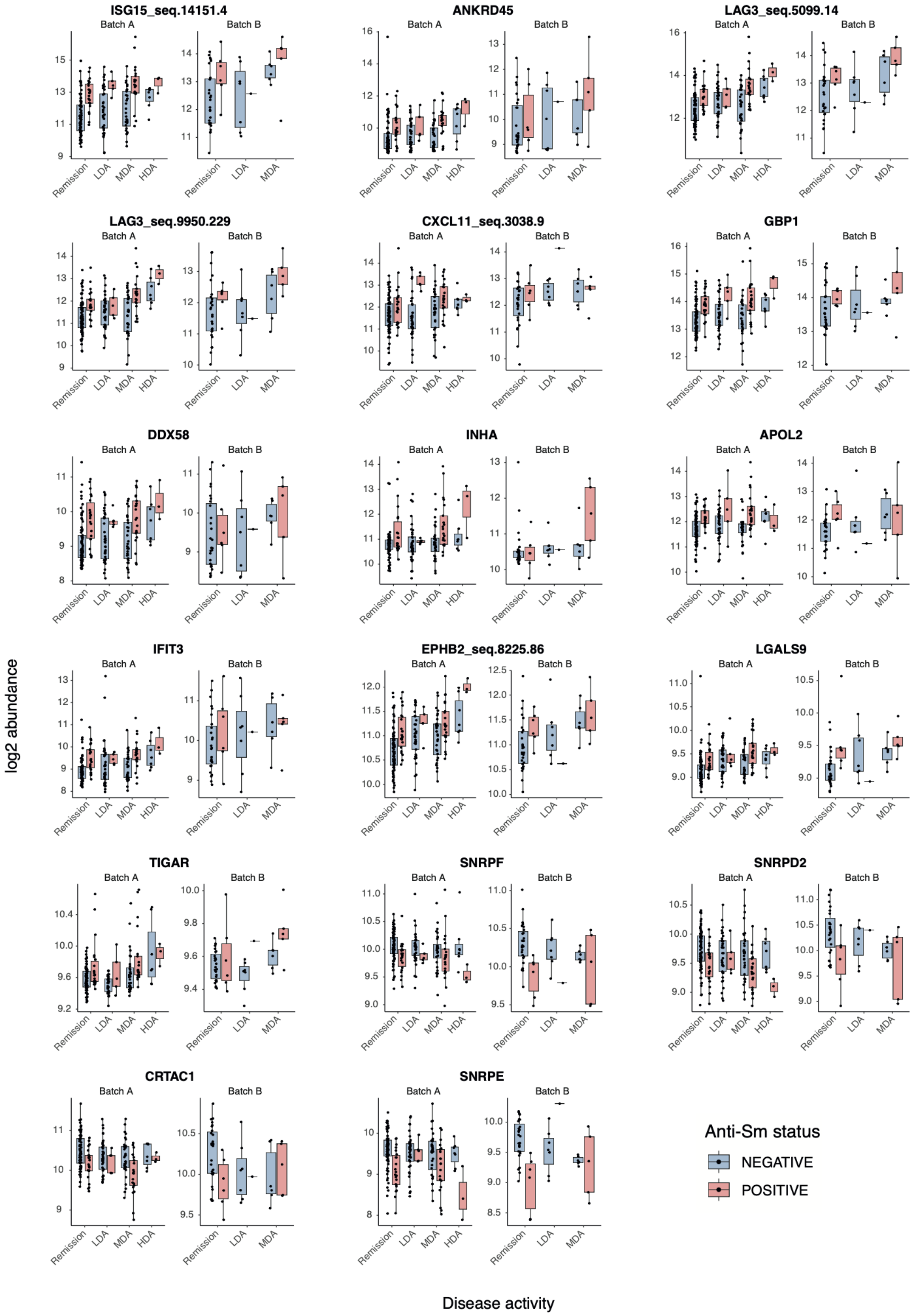


**Supplementary Figure 9. Proteins associated with anti-Sm autoantibodies independent of disease activity.** Boxplots showing the protein levels of anti-Sm-associated proteins (n=17, excluding 3 proteins that were displayed in Figure 4C) stratified by batch and disease activity. For clarity of visualisation, only the proteins associated with anti-Sm positivity at 1% FDR were shown. A full list of anti-Sm associated proteins is provided in Supplementary Data 13.

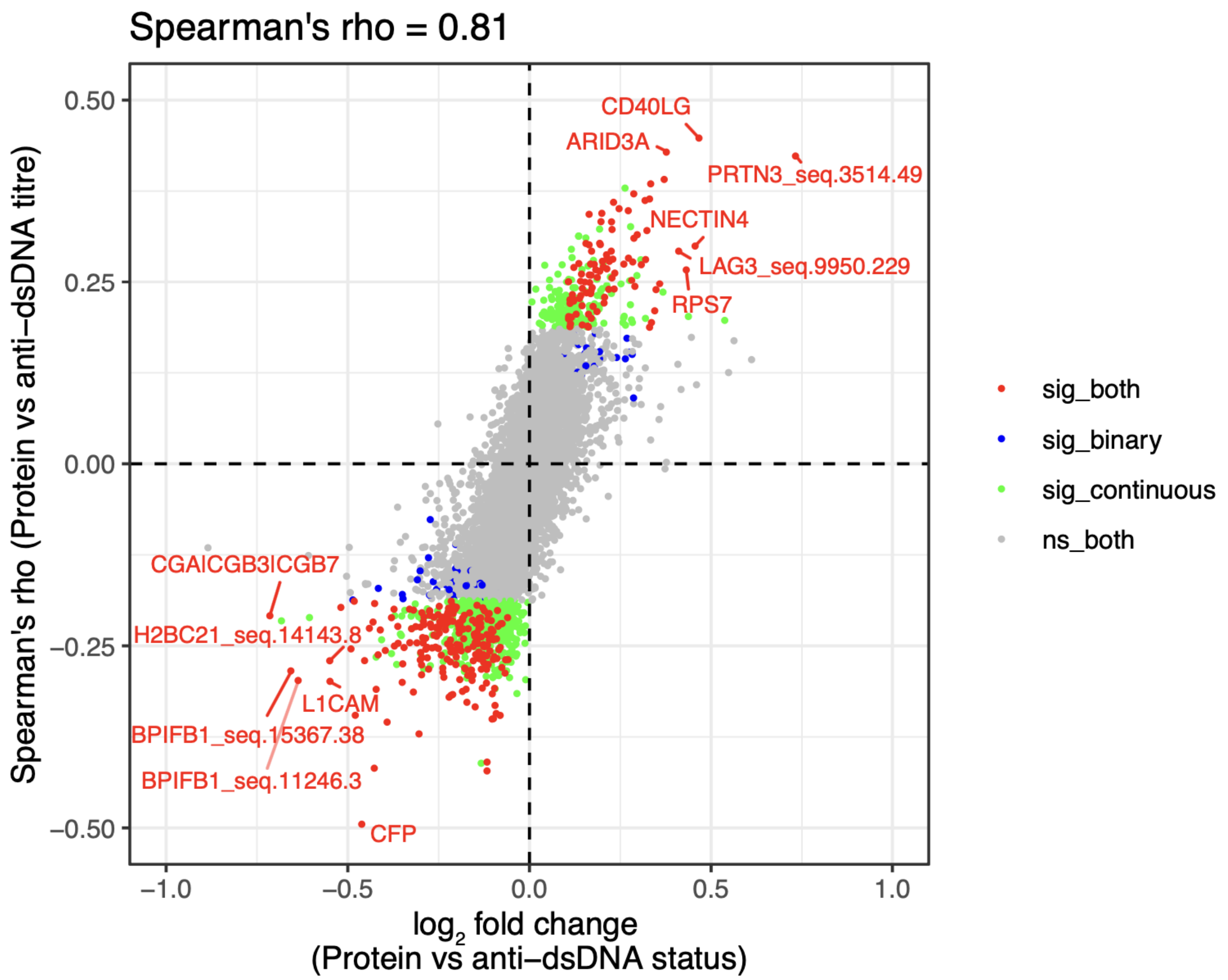


**Supplementary Figure 10. Sensitivity analysis treating anti-dsDNA titre as a continuous variable.** Sensitivity analysis was performed using Spearman's correlation test on protein levels vs anti-dsDNA titre (as a continuous trait) to obtain Spearman's rho (Y-axis) and compared with effect size (adjusted $log_2FC$) obtained from linear regression (X-axis, treating anti-dsDNA status as binary outcome). Protein levels were adjusted for batch, sex, and disease activity in both analyses. Each point represents a protein and is coloured based on their statistical significance - i) not significant (ns) in both models (grey); ii) significant in both models (red); iii) significant only in the linear regression model (blue); or iv) significant only in the Spearman's test (green).

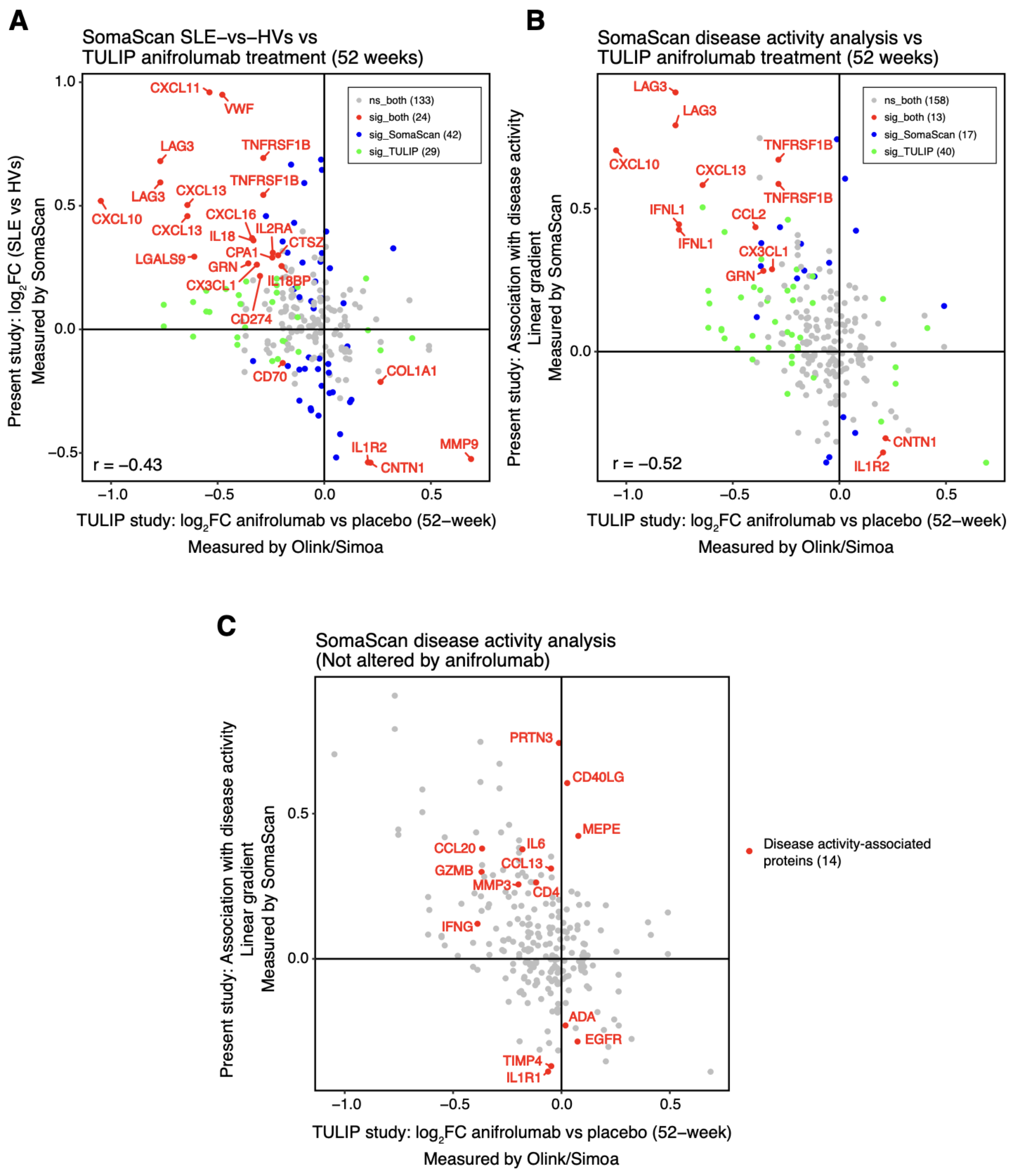


**Supplementary Figure 11. Comparison of proteomic changes in SLE with anifrolumab treatment. A)** Effect sizes ($log_2FC$) from our analysis of SLE vs HVs compared with those from the TULIP-1 anifrolumab trial (52 weeks post-anifrolumab vs placebo), showing an inverse correlation (r = -0.43). Proteins dysregulated (FDR < 0.05) in both datasets are highlighted in red. Grey: proteins ns (not significant) in any dataset. Blue: proteins dysregulated in our analysis only. Green: proteins dysregulated in TULIP-

1 trial only. **B)** Associations with disease activity in our analysis compared with treatment effects in the TULIP-1 study, showing inverse correlation (r = -0.52). Proteins significant in both datasets are highlighted in red. **C)** Proteins associated with disease activity (FDR <0.05) in our study but were not altered by anifrolumab treatment at nominal P <0.05**.**

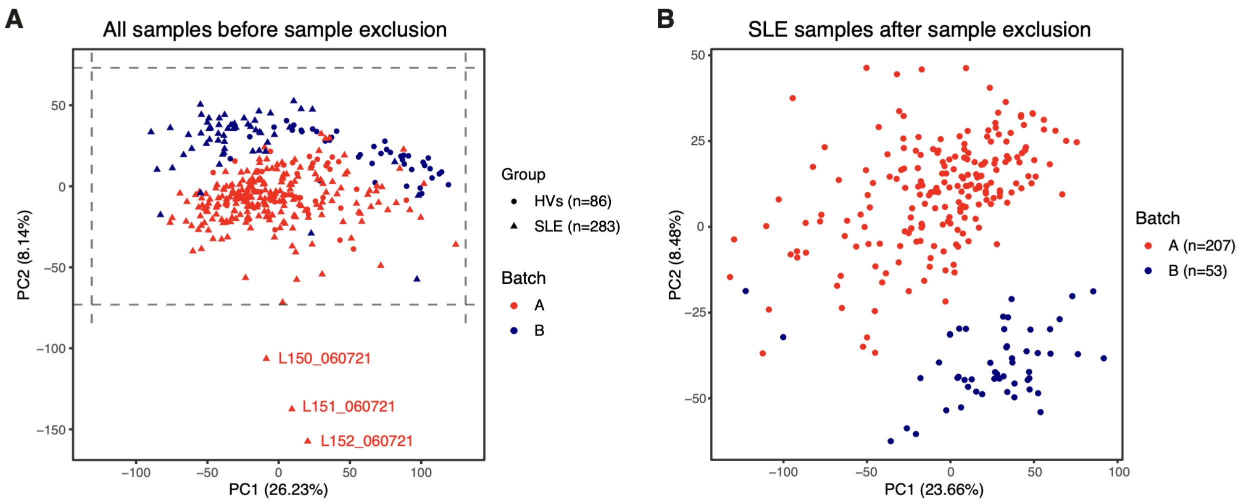


**Supplementary Figure 12. Principal components analysis (PCA) of proteomic data. A**) PCA plot of SLE and HV samples before outlier removal. Each point represents a sample. Batch is indicated by colour and disease status by shape. Dashed lines indicate the range of ±3 SD from the mean of PC1 and PC2. Three outlier samples outside the dashed lines and were processed on the same date are annotated. **B)** PCA plot restricted to SLE samples (after removal of outlier samples). Red: Batch A samples, Blue: Batch B samples.

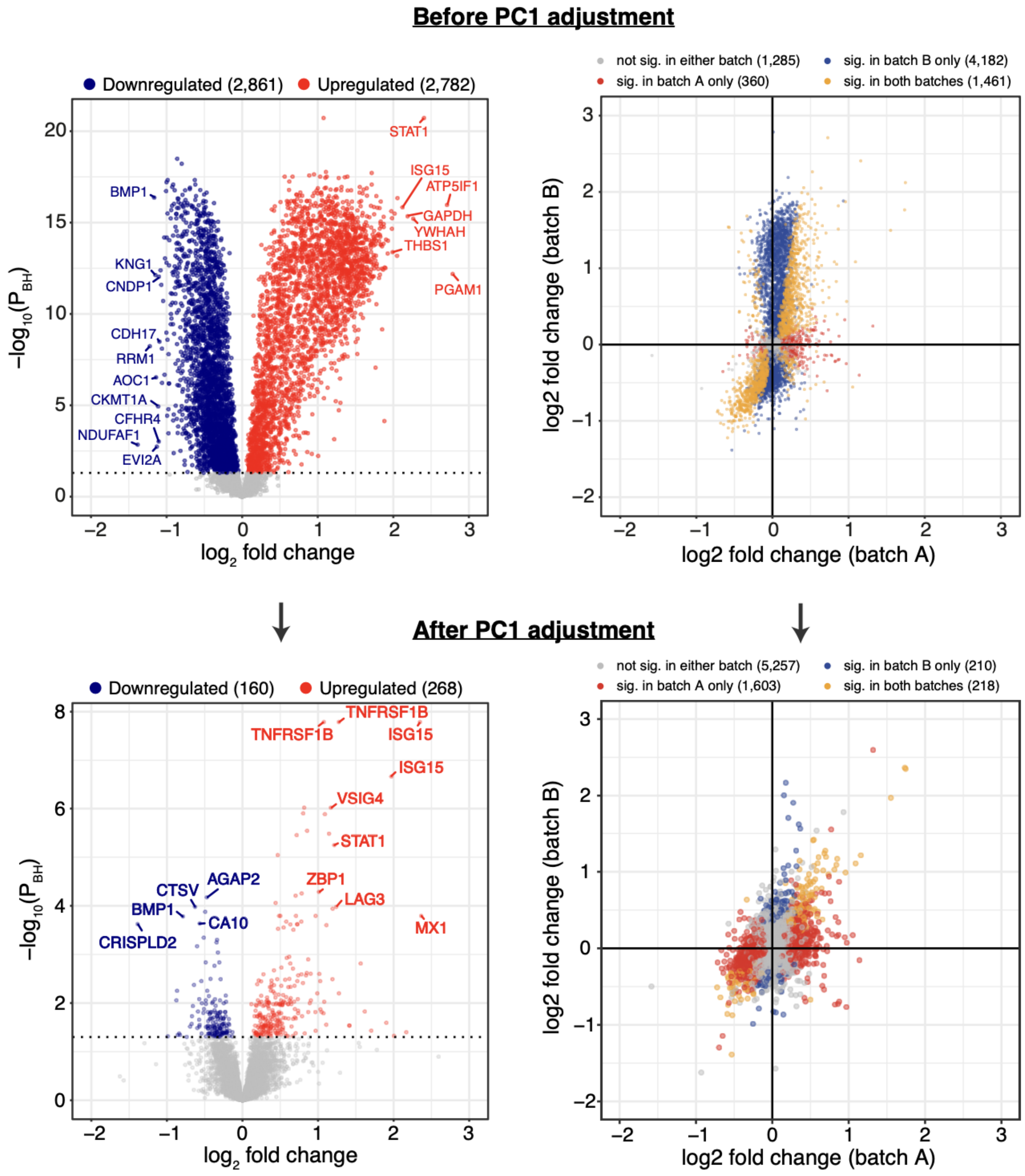


**Supplementary Figure 13. Inflated signals in batch B and mitigation by PC1 adjustment.** An unusually large number of significantly differentially abundant proteins was observed in batch B before PC1 adjustment. The most strongly upregulated protein was PGAM1, a platelet activation marker, suggesting pre-analytical variations. After adjustment for the first PC, inflated signals were substantially reduced. Effect sizes were more consistent across batches.

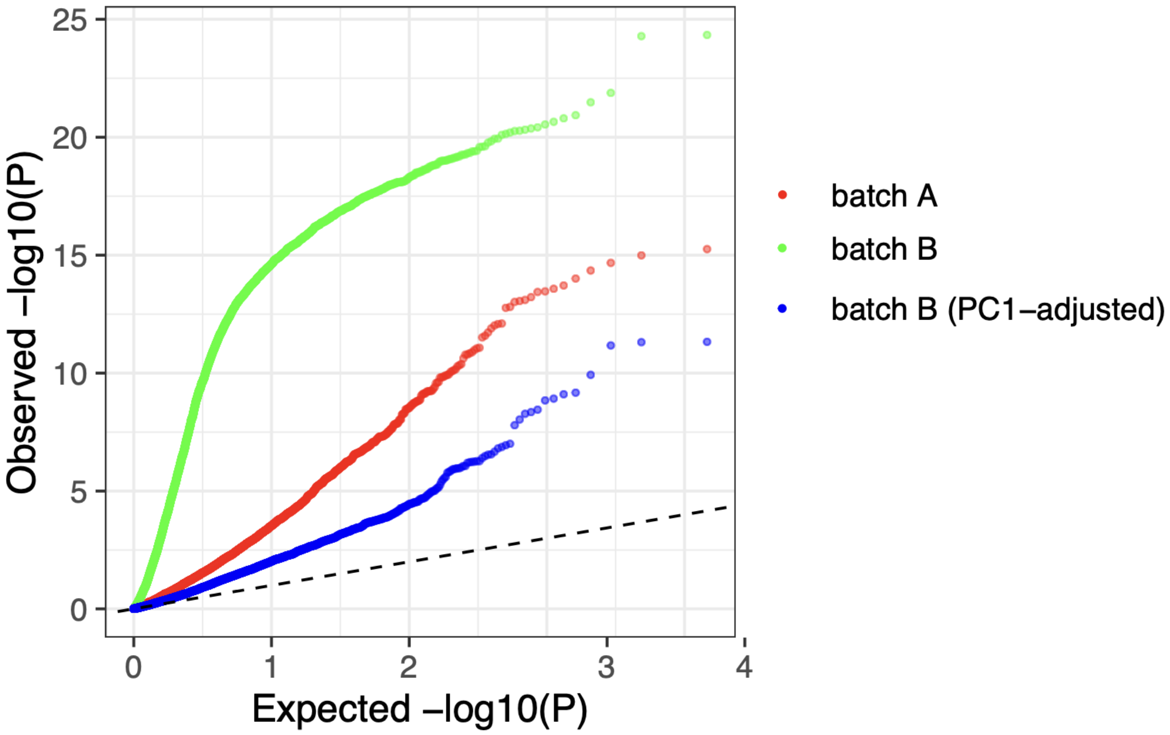


**Supplementary Figure 14. QQ plots of SLE vs HV associations.** Batch B showed strongly inflated test statistics (green) relative to batch A (red), which were largely normalised after PC1 adjustment (blue).

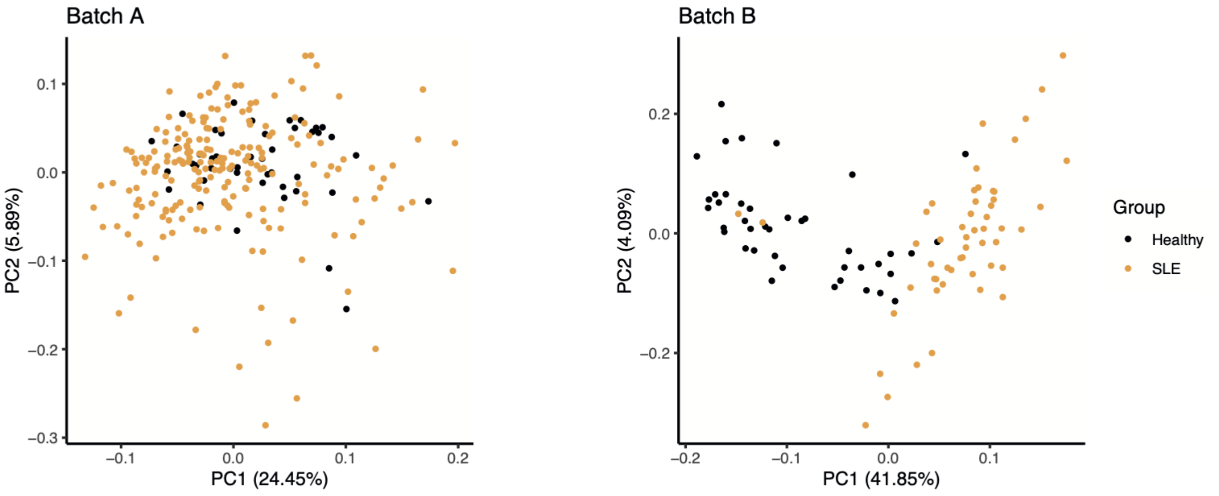


**Supplementary Figure 15. PCA of SLE and healthy samples in two batches of data.** PCA was performed separately on Batch A and Batch B. Each point represents a sample. Orange = SLE, Black = healthy volunteers

#### Titles for Supplementary Data (in Excel file)

**Supplementary Data 1. Distribution of Lupus Nephritis (LN) classes among SLE patients with previous or recent LN.** Contains the number of SLE patients with previous or recent biopsy-confirmed LN in each class.

**Supplementary Data 2. Differential abundance analysis of SLE vs HVs (Batch A).** Contains test statistics and estimates from linear regression model comparing SLE vs HVs in Batch A, and protein information, for all 7,288 proteins.

**Supplementary Data 3. Differential abundance analysis of SLE vs HVs (Batch B).** Contains test statistics and estimates from linear regression model comparing SLE vs HVs in Batch B, and protein information, for all 7,288 proteins.

**Supplementary Data 4. SLE vs HVs differentially abundant proteins in both batch A and batch B.** Contains test statistics and estimates from linear regression model comparing SLE vs HVs in Batch A and Batch B, and protein information, for 215 validated proteins ($P_{BH} < 0.05$).

**Supplementary Data 5. Robust-rank aggregation on pathway terms in SLE vs HVs.** Contains Robust-rank aggregation results and GSEA test statistics of pathways in SLE vs HVs comparisons in Batch A and Batch B.

**Supplementary Data 6. Summary statistics for each protein from disease activity association tests.** Contains test statistics and estimates from linear regression model, and protein information, for all 7,288 proteins tested against disease activity.

**Supplementary Data 7. GSEA results of disease activity association analysis.** Contains effect sizes, test statistics and leading edge for each pathway term obtained from GSEA tested against disease activity associations.

**Supplementary Data 8. Proteins correlated with IFNL1.** Contains correlation coefficient and protein information for 75 proteins correlated with IFNL1 (Pearson's $r > 0.6$, $P_{BH} < 0.05$).

**Supplementary Data 9. Proteins significantly correlated with clinical C3 levels.** Contains correlation coefficient and protein information for 60 proteins correlated with clinically measured C3 levels ($|rho| > 0.3$, $P_{BH} < 0.05$).

**Supplementary Data 10. Proteins significantly correlated with clinical C4 levels.** Contains correlation coefficient and protein information for 62 proteins correlated with clinically measured C4 levels ($|rho| > 0.3$, $P_{BH} < 0.05$).

**Supplementary Data 11. Connectivity of each protein in the red (IFN-associated) module.** Contains connectivity (Spearman's correlation between protein levels and the module's eigenprotein values) and protein information for 35 proteins in the red module.

**Supplementary Data 12. Connectivity of each protein in the blue (renal-associated) module.** Contains connectivity (Spearman's correlation between protein levels and the module's eigenprotein values) and protein information for 611 proteins in the blue module.

**Supplementary Data 13. Proteins associated with anti-Sm positivity before or after adjusting for disease activity.** Contains test statistics and estimates from linear regression model comparing anti-Sm(+) vs anti-Sm(-), and protein information, for proteins that are differentially abundant before or after adjusting for disease activity ($P_{BH} < 0.05$).

**Supplementary Data 14. Proteins associated with anti-dsDNA positivity before or after adjusting for disease activity.** Contains test statistics and estimates from linear regression model comparing anti-dsDNA(+) vs anti-dsDNA(-), and protein information, for proteins that are differentially abundant before or after adjusting for disease activity ($P_{BH} < 0.05$).

**Supplementary Data 15. All proteins measured by SomaScan in this study.** Contains all proteins measured by SomaScan (v4.1) in this study. A total of 7,288 SOMAmers along with their SOMAmer ID, target protein, UniProt ID, Entrez Gene ID, and gene symbol are listed.
